# ImputePGTA: accurate embryo genotyping and polygenic scoring from ultra-low-pass sequencing

**DOI:** 10.1101/2025.11.07.25339763

**Authors:** Jeremiah H. Li, Tobias Wolfram, Ivan Davidson, Justin Schleede, Jennifer Swift, Spencer Moore, David Stern, Michael Christensen, Alexander Strudwick Young

## Abstract

Preimplantation genetic testing (PGT) for polygenic risk (PGT-P) holds great promise for reducing lifetime disease burden, but genotyping embryos remains difficult. PGT for aneuploidy (PGT-A) is a routine test used in over half of *in vitro* fertilization cycles in the United States, typically via ultra-low-pass (ULP) sequencing (∼0.004x) or, less commonly, genotyping arrays. Here we describe an approach that enables accurate embryo genotyping from PGT-A data when combined with estimated parental haplotypes. We develop a Coupled Hidden Markov Model, ImputePGTA, which jointly infers inheritance patterns from parents to offspring as well as phasing errors in parental haplotypes, along with an inference algorithm that scales linearly with the number of embryos. The performance of our approach depends on the phasing of parental haplotypes, which we improve through a method, phaseGrafter, that combines evidence from short and long reads, further enabling imputation of rare variants. We validate our approach through simulations and comparison of embryo genomes reconstructed from real PGT-A data to post-birth whole genome sequencing data. When using long reads for parental phasing, we achieve a dosage correlation of 0.98 with high-quality post-birth genotypes, and a mean absolute difference of 0.11 standard deviations across 17 disease polygenic scores, lower than from imputation of genotyping array data from reference panels. Uncertainty from imputation from ULP PGT-A data with accurate parental phasing results in only a ∼2% attenuation in expected gains from embryo selection for typical embryo cohort sizes. Our approach removes an important technological barrier to using PGT-P and is already facilitating more widespread adoption.

## Introduction

In vitro fertilization (IVF) has expanded rapidly, reaching 432,641 cycles in the United States alone in 2023 and accounting for 2.6% of all births that year ^1^. Demand for pre-implantation genetic testing (PGT) has risen in parallel, with PGT for aneuploidy (PGT-A) performed in 59% of IVF cycles in 2022, up from 14% in 2014^2,3^.

Most human diseases and traits are polygenic and can be predicted to varying degrees using polygenic scores (PGSs) that aggregate the effects of many genetic variants across the genome ^4^. Polygenic embryo testing (PGT-P) uses PGSs to inform implantation decisions and has the potential to reduce human disease burden ^5–8^. PGT-P requires genome-wide embryo genotypes, which remains costly and technically challenging due to the limited DNA that can be obtained from biopsies ^9,10^.

Enabling genome-wide embryo genotyping from the data routinely generated from PGT-A could dramatically lower the barriers to PGT-P. However, PGT-A data is typically ultra-low-pass (ULP) short-read sequencing data, with coverages as low as 0.002x-0.01x ^11,12^. While this is sufficient for detecting large-scale chromosomal abnormalities, standard methods for obtaining reliable variant calls require far higher coverage.

In other contexts, low-pass sequencing combined with reference-panel-based genotype imputation is routinely used to impute genomes from depths as low as *∼*0.1x ^13–16^. However, only variants in a reference panel can be imputed, and accuracy deteriorates rapidly with decreasing allele frequency and for genetic ancestries poorly represented in the reference panel ^17–19^. While these methods are often sufficient for population level association analyses, they are unable to provide accurate individual genotype calls from ULP PGT-A data.

In contrast to population studies, parental genomes are often available for sequencing in the context of IVF. Each embryo’s genome is a mosaic of maternal and paternal haplotypes, implying that genome-wide reconstruction of embryo genotypes could be feasible via identification of the inheritance vectors (IVs) describing this mosaic. Existing approaches span array-based and custom higher-coverage sequencing designs and typically assume near-perfect parental phasing ^9,20–24^. While sophisticated statistical phasing methods have been developed ^25,26^, it is not typically possible to produce chromosome-level haplotype estimates that are sufficiently accurate for comprehensive imputation from ULP data. Switch errors, consecutive heterozygotes that are incorrectly phased relative to each other, result in estimated haplotypes that are *themselves* mosaics of the true haplotypes, making inference of inheritance patterns significantly more challenging.

Here we describe our combined wet-lab and computational pipeline for embryo whole-genome reconstruction (**Figure 1**). To achieve clinical-grade accuracy for embryo genotypes at both common and rare variants, we sequence parents with both short and long reads. We then unify statistical phase estimates generated from short reads (SRs) with read-backed phase estimates from long reads (LRs) using a novel method called phaseGrafter, resulting in chromosome-scale parental haplotypes. We then apply ImputePGTA — a novel Coupled Hidden Markov Model (CHMM) based algorithm — to reconstruct inherited embryo genomes. A key innovation of our approach is computationally efficient joint inference of switch errors in parental haplotypes along with inheritance vectors for all embryos, using an algorithm that scales linearly with the number of embryos. This results in posterior distributions over embryo IVs, genotypes, and PGSs, enabling propagation of imputation uncertainty to embryo trait and disease predictions.

**Figure 1.**
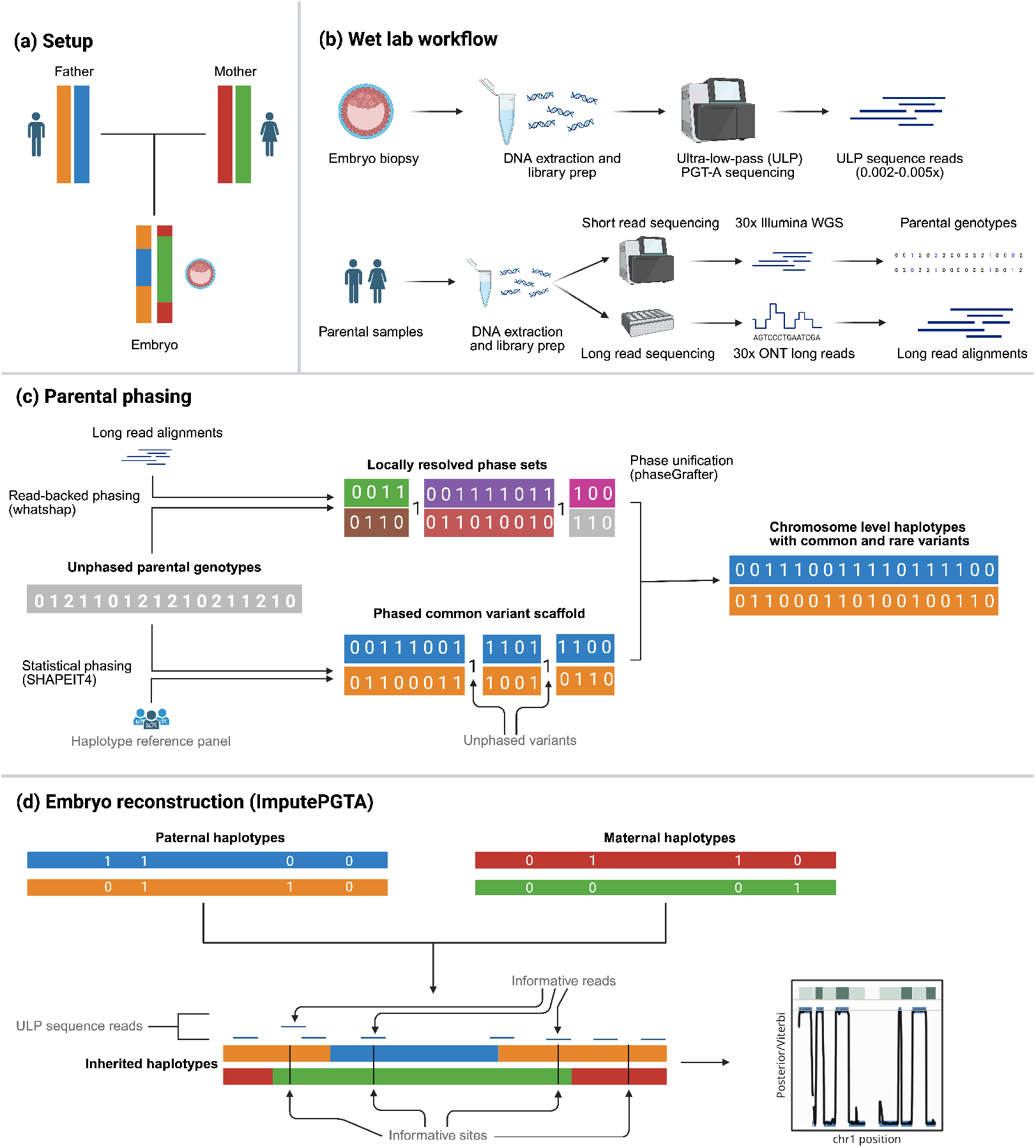
Graphical Abstract: Graphical representation of the workflow used to reconstruct embryo genotypes given sequence data on the parents and ultra-low-pass (ULP) PGT-A data on at least one embryo. **(a)** The experimental setup where we have two parents and at least one embryo. Each parental haplotype is given its own color; each embryo inherits a mosaic of the parental haplotypes. **(b)** The wet lab workflow for the embryos and the parents. A trophectoderm biopsy is taken from each embryo and sequenced at ultra low depth (down to 0.002x). Blood or saliva samples are collected from the parents and sequenced at high depth using both Illumina short reads and Oxford Nanopore long reads. **(c)** Parental haplotypes are estimated using phaseGrafter, our novel dynamic programming algorithm for unifying phase estimates from a common variant scaffold generated from statistical phasing (*e.g*., SHAPEIT4) and independent phase sets generated by whatshap during read-backed phasing using Oxford Nanopore long reads. **(d)** ImputePGTA, our embryo genome reconstruction method, takes as inputs the inferred parental haplotypes and the union of reads overlapping trio-segregating sites (*i.e*., sites where at least one parent is heterozygous) across all embryos. Inheritance and switch vectors are then inferred, producing posterior distributions over parental switch states and offspring genotypes. Posteriors from one of the real PGT-A cases are shown in black; in dark blue are the “true” inheritance vectors inferred from the high-coverage data from the born child. The best-fitting sampled switch-state path from the inferred parental switch vector is shown in green, where a transition from light to darker green indicates a switch.

We validate ImputePGTA in simulations and by comparing genotypes imputed from real PGT-A (or downsampled embryo high coverage whole genome data) data on 33 embryos to high coverage whole genome data from born children (or high coverage embryo whole genome data). When adding long reads to assist in parental phasing, the average dosage correlation for embryos with ULP data increases from 0.960 to 0.980 and the imputed PGS mean absolute error (MAE) relative to the true PGS drops from 0.182 to 0.110 SDs; for embryos with PGT-A array data, dosage correlation increases from 0.992 to 0.994 and PGS MAE drops from 0.067 to 0.049 SDs. As switch errors result in changes in inheritance vectors for all embryos, joint modeling of multiple embryos improves estimation of switch errors and thereby gives more accurate imputed embryo genotypes. For example, for a family with poor parental phasing, the imputed dosage correlation rises from 0.835 with a single embryo to 0.938 with 10 embryos and 0.966 with 25 embryos.

Our results show that accurate genome reconstruction and polygenic scoring, along with posterior uncertainties, can be achieved from routine PGT-A data, removing an important barrier to wider adoption of PGT-P.

## Results

### ImputePGTA for embryo genotyping and polygenic scoring

ImputePGTA (**Figure 1D**) is a novel Coupled Hidden Markov model (CHMM) for imputation of offspring genotypes from ULP sequence data or array data given estimated parental haplotypes (phased genotypes). We focus on the ULP sequence data case as it is the more prevalent and challenging scenario.

Given estimated parental haplotypes and a cohort of *N* embryos, each offspring’s genotype is determined by the haplotypes it copies from (inherits) from each of the parents at each position, which is represented by a binary inheritance vector (IV) for each embryo’s maternally and paternally inherited haplotype. At each genomic position, the inheritance vector takes on values in (0, 0), (0, 1), (1, 0), (1, 1). The first element is the index of the paternal haplotype the embryo copies from at that position, and the second element is the index of the maternal haplotype it copies. The IV changes when there is a cross-over or a switch error in the parental haplotypes. Switch states are similarly represented by a binary switch vector, taking values of 0 (no switch) or 1 (switch), which represent the orientation of the true parental haplotype relative to the estimated haplotype at that position.

ImputePGTA infers a joint posterior distribution over the parental switch vectors and the IVs of all embryos. The key features that make ImputePGTA more effective than previous approaches ^9,24^ are that switch errors in the parental haplotypes are explicitly modeled and inferred, with the switch error rate for each parent first estimated via maximum likelihood, followed by joint inference of the switch vectors and the IVs for all embryos.

The CHMM underlying ImputePGTA scales exponentially in the number of embryos, making exact inference intractable for more than a few embryos (5-10). As the number of euploid embryos resulting from a single IVF cycle can often be higher than this (with upper ranges exceeding 15 for patients under 35, ref. ^8^), we developed a method that enables us to sample from the joint posterior distribution with computations that scale linearly in the number of embryos, while achieving equivalent performance to exact inference.

Posterior samples of the embryo IVs can be used to compute posterior distributions of PGSs for embryos, enabling the propagation of uncertainty to predictions of disease risks in PGT-P (**Methods**). To illustrate this in a realistic setting, we used PGSs for 17 diseases from a recently published manuscript covering several cancers, metabolic and cardiovascular diseases, Alzheimer’s disease, multiple sclerosis, inflammatory and autoimmune diseases, glaucoma, and osteoporosis ^7^ (**Methods**).

We validated the performance of ImputePGTA in simulations and on downsampled 30x WGS data from a cohort of 25 real embryos. Three aspects we aimed to characterize were the effects of (1) switch errors in the parental haplotypes as described by *λ*, the rate of single switch errors per cM in each parent (**Methods**), (2) embryo cohort size, and (3) embryo sequencing data coverage. We characterize genotype imputation accuracy in terms of genome-wide dosage correlation, and the accuracy of PGS estimated from the imputed genotypes in terms of mean absolute error (MAE) from the true PGS.

### Simulated offspring

We simulated 20 offspring from gold-standard parental haplotypes for two parents in the Platinum Pedigree ^27^ (**Methods**). To characterize the effect of *λ*, embryo coverage, and cohort size, we induced switch errors in both sets of parental haplotypes with *λ* ranging from 0cM^*−*1^to 1cM^*−*1^(covering the typical range observed in real data), simulated sequencing reads on the offspring at coverages spanning those typical for a PGT-A assay (0.002x, 0.004x, 0.01x), and performed inference at various cohort sizes.

ImputePGTA first infers *λ* for each parent. We observed that our estimates of *λ* were accurate and well-calibrated (**Supplementary Figure S1**). Joint inference of the parental switch vectors and the embryo IVs is then performed, and embryo genotypes are inferred from the resulting marginal posterior distributions. We compared the imputed genotypes to the known genotypes of the simulated offspring at trio-segregating (TS) sites, defined as sites where at least one parent is heterozygous.

### Dosage correlation

Overall, we observed very high accuracies when *λ* is low, with performance degrading as *λ* increases (**Figure 2A, Supplementary Table 1**). Performance increases as the cohort size increases, with substantial gains even with just one additional embryo (*n* = 2), and diminishing returns for additional embryos as the cohort size increases. At 0.002x, the genome-wide correlation between the posterior mean genotype (dosage) and true genotypes ranged from 0.990 at *λ* = 0 to 0.946 at *λ* = 1 for joint inference across a cohort of 20 embryos (**Supplementary Table 1**). When embryos are imputed alone (*n* = 1), dosage correlation at *λ* = 0 remains high (0.990) but drops substantially at *λ* = 1 (0.748), demonstrating the benefit of joint inference.

**Table 1.** Details of the families for which we obtained real embryo data. We obtained real PGT-A data for 6 families from both implanted (focal) and non-implanted embryos and high-coverage WGS data on the corresponding born child(ren) (FAM1-FAM6). For FAM7 (the “downsampled dataset”), we obtained high-coverage WGS data from 25 embryos but no data from a born child; the high-coverage WGS data was treated as the truth data for validation. The “Focal Embryos” column indicates the number of born children (or in the case of FAM7, the total number of embryos). The data modality of the PGT-A assay used during the IVF process (either sequencing or genotyping array) is noted in the “Embryo Data Type” column — all ULP sequencing data had coverage *∼*0.004x; the WGS data had coverage *∼*30x. The “Total Embryos” column denotes the total number of embryos from each family used in joint inference. The “Data source” column indicates the provider that originally performed and delivered the results for the PGT-A assay. The two arrays used were the Illumina HumanCytoSNP-12 and Axiom UKB Array. The “Grandparental data” columns indicate which grandparent(s) were sequenced in this study and used for parental phasing. The genetic ancestries of the parents are noted in the last two columns.

| Family ID | Focal Embryos | Total Embryos | Embryo Data Type | Data Source | Grandparental Data | Paternal Ancestry | Maternal Ancestry |
| --- | --- | --- | --- | --- | --- | --- | --- |
| FAM1 | 1 | 4 | ULP | IGENOMIX | Both grandmothers | EUR | EUR |
| FAM2 | 1 | 2 | ULP | IGENOMIX | None | EUR | EUR |
| FAM3 | 2 | 12 | Array (Affymetrix) | Genomic Prediction | None | EUR | EUR |
| FAM4 | 2 | 21 | Array (Illumina) | Natera | None | EUR | EUR |
| FAM5 | 1 | 9 | ULP | IGENOMIX | None | Admixed (EUR, AFR, AMR) | EUR |
| FAM6 | 1 | 14 | Array (Illumina) | Natera | None | EUR | East Asian |
| FAM7 | 25 | 25 | 30x WGS (downsampled) | Orchid | All grandparents | Admixed (SAS, EUR) | Admixed (EUR, EAS) |

**Figure 2.**
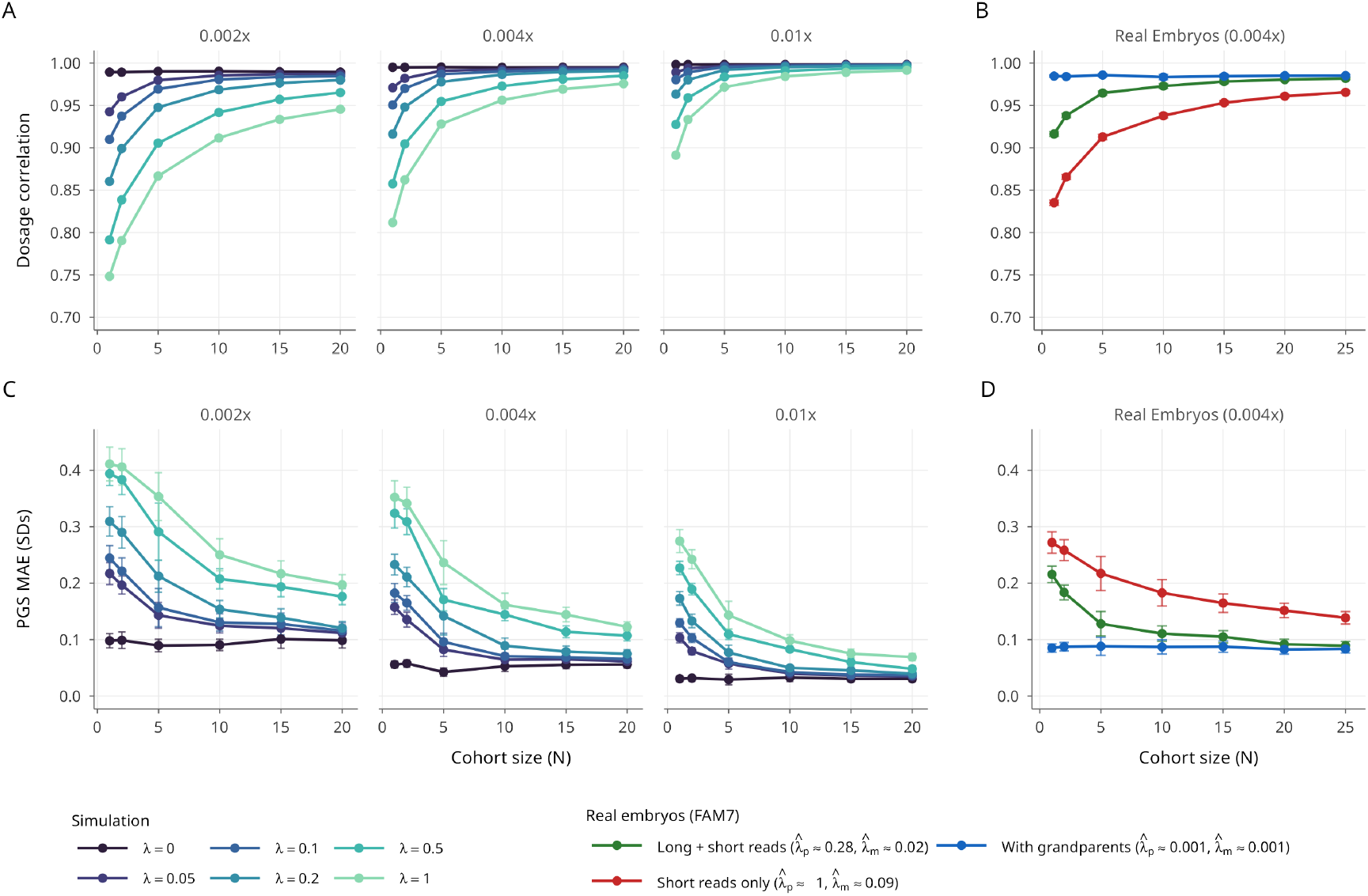
Simulations and downsampled data. **(a)** Imputation accuracy in terms of mean genome-wide correlation between imputed genotype dosages (dosage correlation) and true genotypes at trio-segregating sites across a range of sequence depth (coverage) and switch-error rates (*λ*) in the parental haplotypes for up to 20 simulated offspring of the Platinum Pedigree parents. The points for cohort size *n* = 1 represents the mean over 20 independently imputed embryos, and those for *n* = 2 represent the mean over the same 20 embryos resulting from 10 runs of imputation where each run corresponds with non-overlapping subsets of 2 of the 20 embryos. For all other cohort sizes, they represent the mean over a single *n*-embryo subset of the 20 embryos. **(b)** The same for the 25 real downsampled embryos from FAM7 under three parental phasing qualities (**Results**). The points for *n* = 1, 2 cohort sizes represent means generated in the same manner as above. **(c)** Mean absolute error between the posterior mean PGS and true PGS in units of standard deviations of the PGS. 95% confidence intervals are shown as whiskers. Averages are across the simulated offspring as in (a) and 17 disease PGSs at each (*λ*, coverage, *n*) combination. **(d)** the same as for (c) but for the 25 real downsampled embryos.

### Polygenic score accuracy

For each embryo, we generated 500 posterior samples of their IVs, and scored them for the 17 disease PGSs, resulting in posterior distributions for each PGS for each sample. **Figure 2C** shows the MAE (in units of PGS standard deviations) of the posterior mean PGS estimate relative to the true PGS. At a coverage of 0.004x, the MAE was 0.061 when *λ* = 0.05, rising to 0.123 when *λ* = 1 for joint inference across a cohort of 20 embryos. When embryos are imputed individually, the MAE was 0.158 at *λ* = 0.05 and 0.352 at *λ* = 1 (**Supplementary Table 3**). We computed 95% equal-tailed credible intervals (CIs) of the PGS distributions and found them to be well calibrated, with an average empirical coverage of 95.1% across the parameter space (**Supplementary Figure S2**).

These results indicate that parental phasing quality, embryo cohort size, and embryo sequencing coverage are the three key parameters that control the expected imputation performance and quality of PGS estimates — if phasing performance is expected to be low, then increasing embryo coverage can compensate for this limitation. Similarly, if only a small number of embryos are available, parental phasing quality and embryo coverage have greater relative importance.

### Effect of imputation uncertainty on selection efficacy

The expected gain from embryo screening for disease traits scales with within-family PGS variation on the liability scale (see **Methods** for details). Posterior uncertainty shrinks imputed PGS toward the family mean, reducing between-embryo variation and thus selection gains. The attenuation factor (**Methods**) due to imperfect imputation is

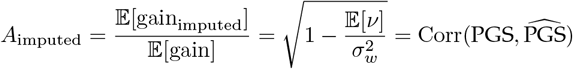

Where 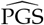 is the posterior mean PGS, 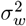 is the within-family PGS variance, and 𝔼[*ν*] is the mean PGS posterior variance across the imputed embryos.

To validate this, we simulated 100 batches of 5 offspring (the median number of euploid embryos resulting from a single round of IVF for a 30 year old woman ^8^) from the Platinum Pedigree parents, imputed from simulated 0.004x coverage read data under parental switch error rates of *λ* = 0.05cM^*−*1^, 0.125cM^*−*1^, or 1cM^*−*1^, and computed gains for 17 traits (**Methods**). Empirical gains from screening on imputed PGSs closely matched the theoretical prediction (**Figure 3**), with negligible attenuation at *λ* = 0.05cM^*−*1^and *λ* = 0.125cM^*−*1^(*A*_imputed_ = 0.98 and 0.97) and a larger drop at *λ* = 1cM^*−*1^(*A*_imputed_ = 0.85), again highlighting the importance of accurate parental phasing, especially in the absence of a large number of embryos. Gains on true PGSs ranged from 0.46 to 0.92 SDs across traits (**Supplementary Table 10**), showing that the expected efficacy of selection can vary substantially from PGS to PGS within-families due to differences in within-family variance (Methods), with corresponding absolute and relative risk reductions shown in **Supplementary Figures S3 – S4**.

**Figure 3.**
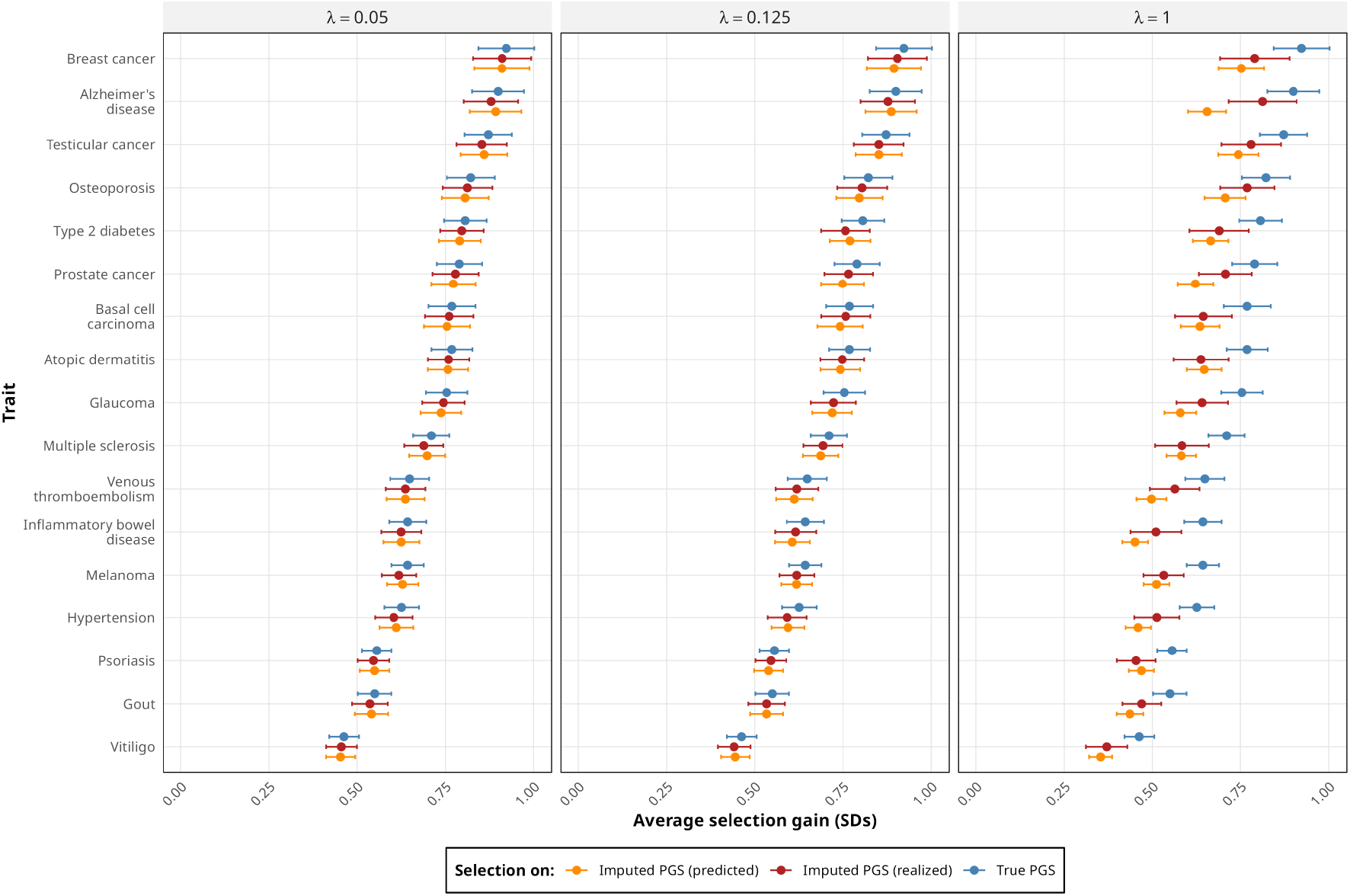
Attenuation of predicted gains due to selection on imputed PGS. The *x*-axis shows the average gain (95% CIs), defined as the absolute difference between the selected embryo’s PGS and average PGS in units of PGS SDs from 100 instances of 5-embryo families with the same parents. The panels show results for three values of *λ*, the switch-error rate in parental haplotypes: the value based on approximate empirical results for parents phased with short reads and long reads, (*λ* = 0.05cM^*−*1^), short reads only (*λ* = 0.125cM^*−*1^), and the highest value analyzed in the *in silico* experiments (*λ* = 1cM^*−*1^). For each panel, three results are shown: the first two are the mean gain from two selection strategies: (a) selection based on true PGS values (blue), and (b) selection based on the imputed PGS values (red). The third is the theoretical gain predicted by the posterior variances (orange). The difference between the red and blue data points reflects the actual decrease in expected gains (relative to true PGS values) resulting from imputation uncertainty.

### Application to real embryo data in seven families

To validate ImputePGTA in real world settings, we conducted a study of seven families who underwent IVF where real PGT-A data was generated (**Methods**). For six of these families, this was followed by successful implantation, pregnancy, and live birth of at least one offspring (**Table 1**). This enabled us to compare imputed embryo genotypes and PGSs from real PGT-A data to high-quality whole-genome sequence data from the born children. We analyzed three born children originally assayed using an ULP sequence-based PGT-A test and five originally assayed using genotyping arrays. For the last family (FAM7), we obtained high-coverage WGS data from 25 embryos but no data from a born child; instead, we used the high-coverage WGS data as truth data for validation. Joint inference used all embryo data available per family, including unimplanted embryos, while accuracy metrics are reported for the focal embryos with truth data (**Table 2**).

**Table 2.**
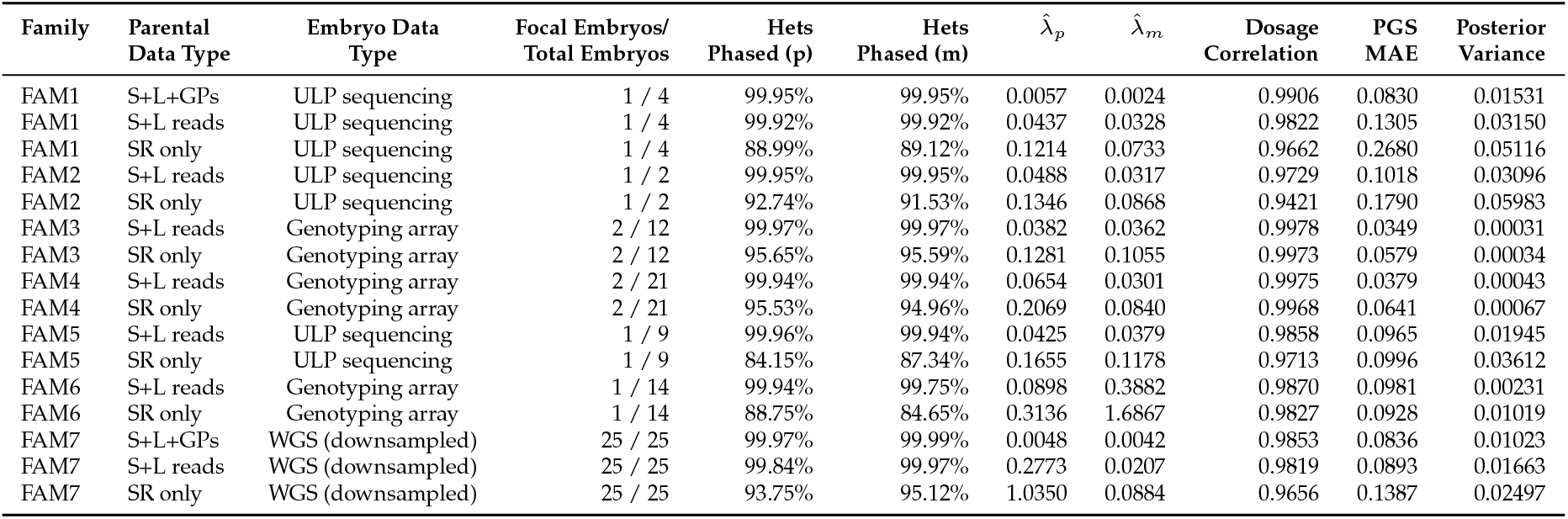
Parental phasing and imputation performance. The percentage of heterozygotes phased and the estimated switch error rates 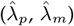 for each parent—paternal (p) and maternal (m)—in each family under the various phasing scenarios is shown, alongside genotype and PGS accuracy. Embryo data type was either ULP sequence data (*∼*0.004x), genotyping array data (Table 1), or WGS data downsampled to 0.004x (FAM7 only). “Focal Embryos / Total Embryos” denotes, respectively, the number of imputed embryos for which accuracy metrics were calculated and the total number of embryos in each family used for joint inference. Imputation accuracy is measured by the dosage correlation between imputed results and true genotypes at trio-segregating (TS) sites. PGS accuracy is described by the mean absolute error (MAE) of the posterior mean estimates in SD units across 17 disease PGSs; posterior variance is averaged across traits and focal samples.

For the three embryos with ULP sequence data, the reads were single-end IonTorrent short reads with an average read length of 112bp and an average coverage of 0.0046x (range of 0.0043-0.0048x) (**Methods**). The embryos with array data were genotyped using two different chips (**Table 1**). Each pair of parents was sequenced with both Illumina SRs and Oxford Nanopore long reads in order to genotype and phase all SNPs and small indels in the parents, including rare variants, using our pipeline (**Figure 1**) (**Methods**). The born children were sequenced at high depth (average 37.1x) using Illumina SRs to determine their “true” genotype.

For FAM1, we also sequenced both grandmothers of the born child using SRs; for FAM7, we sequenced all four grandparents (**Table 1**). Grandparental data enables phase resolution at a subset of heterozygotes in each parent by applying Mendelian inheritance rules, further improving parental phasing.

### Whole-genome parental phasing using phaseGrafter

The accuracy of ImputePGTA depends significantly on the phasing quality of the parental haplotypes. While statistical phasing can be locally accurate, only variants also present in the reference panel can be phased. Sequencing the parents with long reads enables the phasing of these omitted rare variants ^28^, while also improving statistical phase estimates by providing prior information ^29^ (**Table 2**). To unify phase information from these orthogonal sources, we developed a phasing pipeline that is able to integrate information from statistical phasing, read-backed phasing from long reads, and grandparental data, to produce consistent haplotypes using a novel dynamic programming algorithm called phaseGrafter (**Methods**). The results of this pipeline are chromosome-level haplotypes for the parents that span both common and rare variants (**Methods**). To highlight the value of long reads, we phased all seven sets of parents once using both short and long read data (the “base case”), and once using only SR data.

Table 2 shows the percentage of parental heterozygotes phased and the inferred *λ* for each parent under each scenario. In the base case, we phased an average of 99.93% of heterozygotes across all parents, increasing to 99.97% with grandparental data, while only an average of 91.28% were phaseable using only SRs. The inclusion of long reads resulted in an average 3.3-fold decrease in *λ* compared to using SRs only, and using grandparental data further decreased the inferred switch error rate to values approaching zero for FAM1 and FAM7 (**Table 2, Figure 4**).

**Figure 4.**
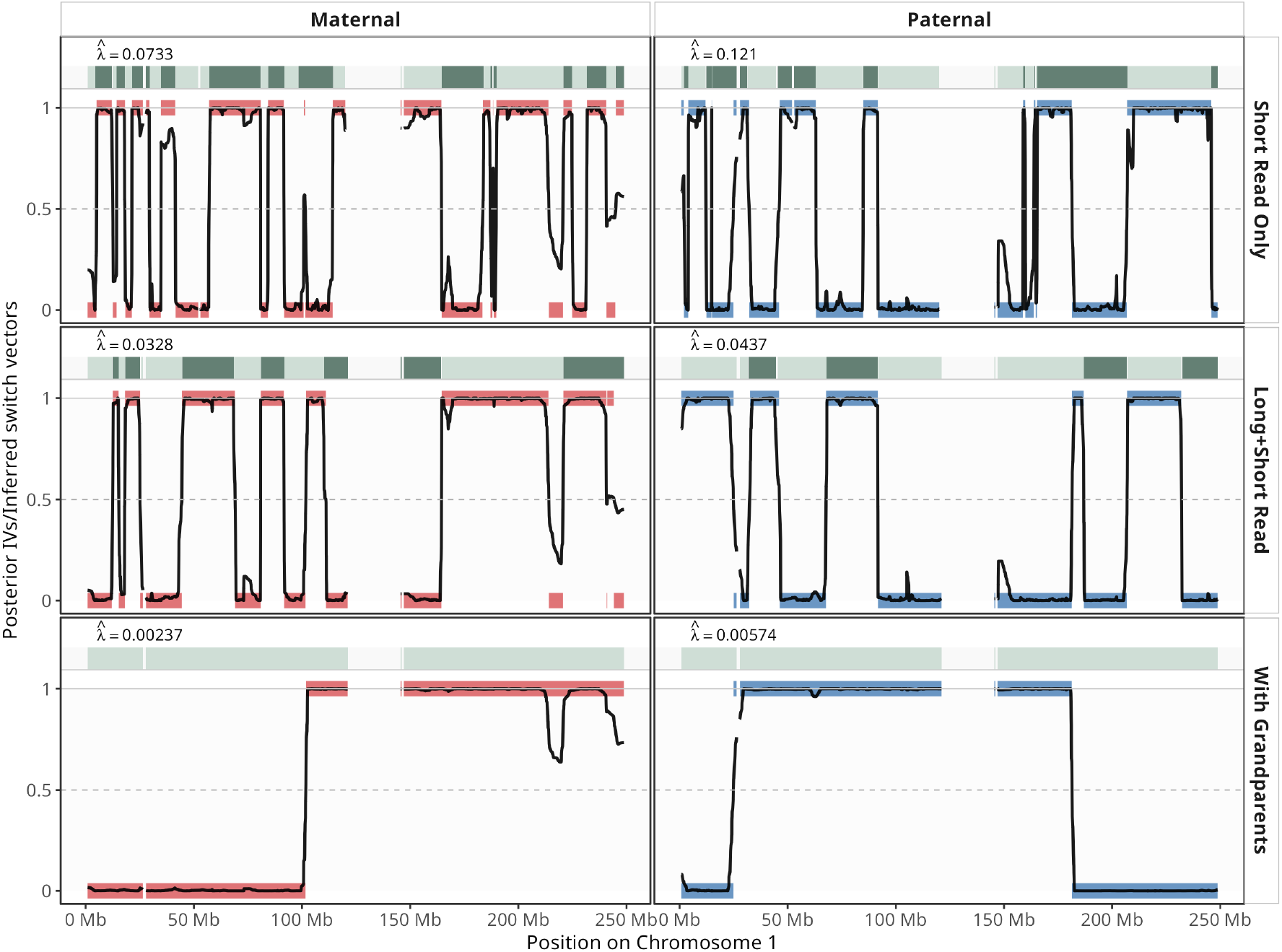
Inferred inheritance vectors and switch errors for FAM1 given different parental phasing strategies. For the embryo from FAM1 with ULP PGT-A data, marginal posterior probabilities referencing the *estimated* parental haplotypes on chromosome 1 are displayed in black. Probabilities at 0 and 1 indicate inheritance of one or the other of the parents’ estimated haplotypes. A transition from 0 to 1 (or vice versa) indicates an inferred crossover event or switch error. The green bars at the top of each panel show the best-fitting sampled path for the inferred parental switch states, with transitions between dark and light green indicating the locations of the inferred switch errors. The thick colored bars in the marginal plots show the rounded posterior probabilities obtained from running ImputePGTA on the born child’s genotypes obtained from high-coverage WGS and reflect a “best-case” estimate for estimation of the true IVs. Top to bottom: phasing parents using short reads only; short and long reads, but without grandparental data; short reads, long reads, and grandparental data. The inferred values of *λ* are shown in each pane. The effect of lower values of *λ* (more accurate phasing) can be seen as fewer switches in IVs needing to be inferred accurately from sparse PGT-A data. The best-fitting sampled switch-state paths track the IV switches closely, though not all switch errors are detected. For the phasing with grandparental data, no switch errors were inferred, implying that the switches in the IVs for that scenario correspond with true recombination events. Gaps on the *x*-axis reflect regions of at least 1Mb where no variants were called in the parents (*e.g*., in centromeric regions).

### Imputation of embryo genotypes

Using the same metrics as the *in silico* experiments, we compared posterior embryo genotype estimates to high-coverage whole genome sequencing genotypes from born children (**Table 2**). For the three families with ULP sequence data and born-child data (FAM1, FAM2, FAM5), the average dosage correlation at trio-segregating (TS) sites was 0.980 in the base case and 0.960 when using SRs (**Supplementary Table 9**). For the three families with embryo array data (FAM3, FAM4, FAM6), we observed even higher imputation accuracy: at TS sites, the average dosage correlation was 0.994 in the base case and 0.992 with SRs only.

For FAM7, where we lacked a born child but had 30x WGS data for all 25 embryos, imputation accuracy resulting from joint inference across all embryos from the downsampled 0.004x data was similar at TS sites, with an average dosage correlation 0.982 in the base case (**Table 2, Supplementary Table 9**). Using grandparental data further increased the dosage correlation (**Table 2, Supplementary Table 5**).

As expected, joint inference accuracy scaled with cohort size: with SR phasing only, the mean dosage correlation increased from 0.835 at *n* = 1 to 0.966 at *n* = 25, while under the best phasing the improvement with *n* was negligible (**Figure 2, Supplementary Table 2**).

Figure 4 compares ImputePGTA marginal posterior inheritance vector probabilities from FAM1 to those inferred from the high-coverage post-birth data (**Supplementary Figures S6 – S8** shows all autosomes), alongside the inferred switch errors in the estimated parental haplotypes. The effect of phasing quality is readily apparent: higher values of 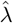 correspond to a more frequent switching of IVs, leading to greater difficulty in inferring the switches, and resulting in regions where the posterior probabilities are intermediate, most visible in the transitions in the top panel of **Figure 4**. The more often the probability is close to 0.5, the larger the posterior variance of the downstream PGS estimates and the less effective selection becomes (**Methods and Figure 3**). The best-fitting sampled paths for the parental switch states track the IV switches closely, indicating that ImputePGTA is able to accurately infer the locations of switch errors. In the case where grandparental data is used, no switch errors are inferred, indicating that the inferred switches in the IVs represent true recombination events in the embryo.

### Inheritance of a rare pathogenic variant

To illustrate the utility of long-read sequencing of parents for imputation of rare variants in embryos, we identified that the father and child in FAM1 carry a rare pathogenic frameshift variant in *VPS13C* (NG_- 027782.1:g.145304dup, rs1315150327), resulting from a single thymine insertion at chr15:61920162 (GRCh38) (**Methods**). This variant leads to a frameshift at codon 2461, introducing a premature termination codon and is associated with autosomal recessive early-onset Parkinson disease ^30^. The variant has an allele frequency of 4.4*×*10^*−*6^ in gnomAD v4.1^31^. In the UKB 200k WGS phased release, only two copies of this allele are observed — at this allele count, statistical phasing performs poorly ^26^. The inclusion of long reads on the parents, however, enables us to correctly impute this variant in the embryo with posterior probability *>* 0.995.

### Polygenic scores

We sampled 500 posterior IVs for each embryo to obtain posterior distributions for 17 disease PGS, and computed the MAE of the posterior mean PGSs relative to the PGSs calculated from the ground truth for each embryo (**Methods, Supplementary Figure S5, Table 2**). Generally, the born child’s PGS overlapped with regions of high posterior density, with some exceptions especially when the posterior uncertainty is very low; in these cases, absolute error also tended to be very low (*e.g*. inflammatory bowel disease in the array cases) (**Figure 5**).

**Figure 5.**
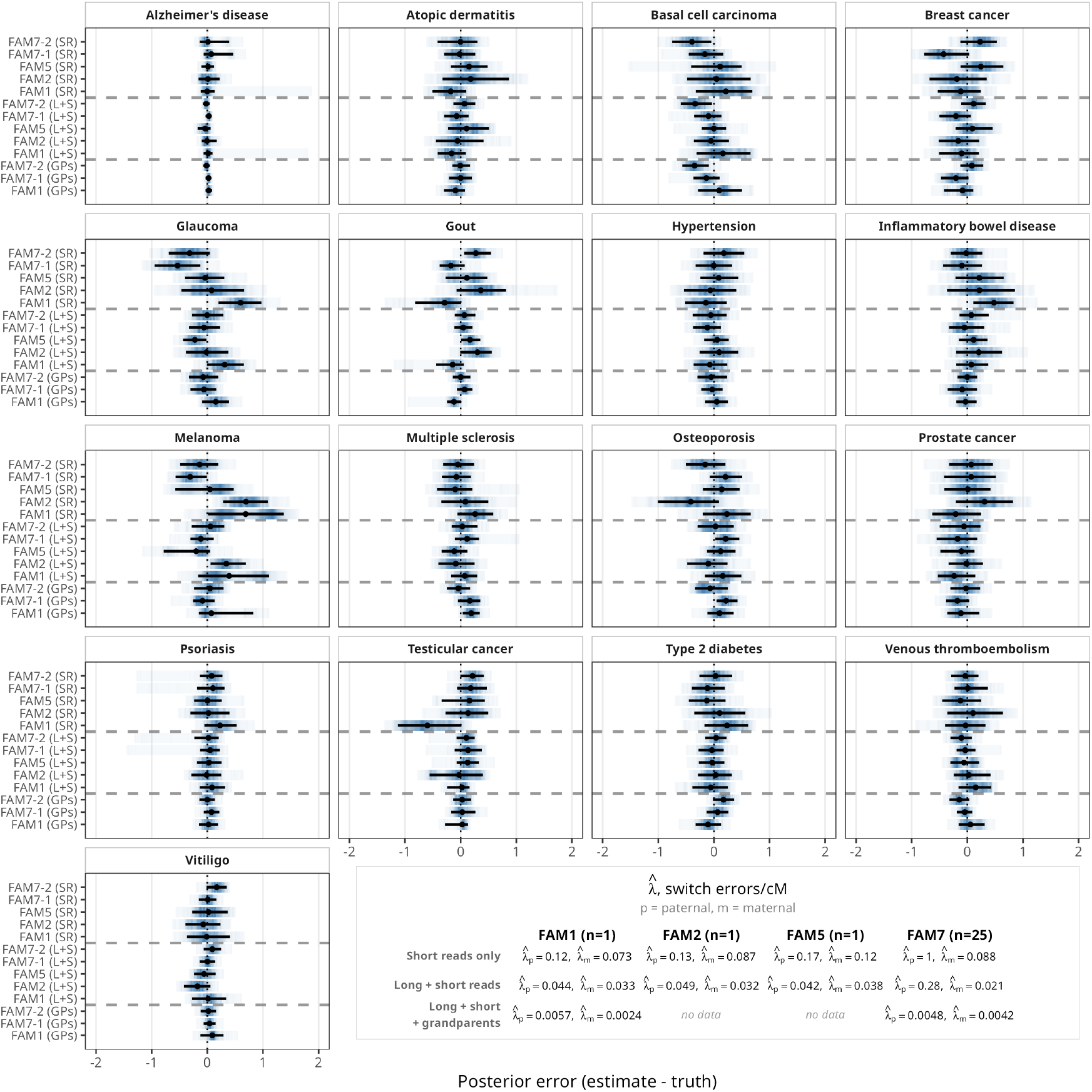
Polygenic score estimates for embryos with ultra-low-pass data. For the families with ULP PGT-A data (FAM1, FAM2, FAM5), we give the difference between the posterior PGS estimates and the PGS calculated from high-coverage data in units of PGS standard deviations across 17 disease PGSs. The black points show the posterior mean PGS, the whiskers denote the 95% equal-tailed credible interval, and the posterior densities are given by the intensity of shading. (SR) indicates parental haplotypes were produced from short-reads only using statistical phasing. (L+S) indicates haplotypes produced from short and long reads. For FAM1 and FAM7, we also give results for haplotypes produced using short and long reads as well as the available grandparental data. Results for FAM7 are also shown though we include only two representative embryos of the 25 for visual clarity. The estimated *λ* values for each parent under each phasing scenario are also given in the inset.

Posterior densities are generally non-Normal, highlighting the importance of sampling PGS posteriors for producing phenotype predictions that properly account for uncertainty in PGS values without making distributional assumptions (**Methods**).

For the three ULP families with born-child validation (FAM1, FAM2, FAM5), the average MAE was 0.110 SDs in the base case and 0.182 SDs with short reads only. For FAM7 (25 downsampled embryos), the average MAE was 0.089 SDs in the base case and 0.139 SDs with short reads only, reflecting the effect of joint inference over a large number of embryos. With grandparental data, the MAE decreased to 0.083 SDs for FAM1 and 0.084 SDs for FAM7 (**Supplementary Table 4**). In the base case, 94.1% (ULP families) and 92.7% (FAM7) of the estimated 95% CIs covered the true value of the PGS, confirming that our posteriors are well-calibrated. (**Supplementary Table 13**).

The PGS estimates for embryos with genotyping array data are even more accurate, achieving an average MAE of 0.05 SDs (0.07 SDs when using only parental short reads). To contextualize this, we also calculated the MAE of PGSs for the embryo array data after applying reference-based imputation instead of ImputePGTA, using a reference panel of 200k haplotypes (**Supplementary Figure S5 Methods**). The average reference-imputed MAE was 0.276 and 0.174 for the embryos genotyped using the Illumina HumanCytoSNP-12 and Axiom UKB Array respectively — higher than the MAE from our method applied to the embryo array data under all phasing scenarios, and similar to results for the ULP embryo data when using SRs only. This suggests that our PGS estimates from ULP data achieve comparable or superior accuracy to those derived from standard approaches in human genetics research ^19,32,33^, while also providing posterior uncertainties. The largest estimated mean posterior variance across traits was for FAM2’s embryo imputed with parental SRs only: assuming a within-family PGS variance of 0.5, this corresponds to an attenuation factor of 0.88. For the family with the lowest mean posterior variance (FAM3), this factor is 0.999, indicating effectively no loss in selection power. However, these estimates are likely slightly over-optimistic due to imperfect calibration of posterior uncertainty in real data.

## Discussion

ULP PGT-A sequencing is now routine in IVF. Here we show that when parental genomes are available, this ULP data can be used to obtain highly accurate embryo genotypes, with accuracy depending on the number of available embryos, the sequencing coverage, and the phasing quality of parental haplotypes. For trio-segregating sites, our approach, ImputePGTA, achieves average genotype dosage correlations exceeding 0.94 under all tested scenarios in real ULP PGT-A data, rising to over 0.97 when using long reads. For PGT-A genotyping array data, which is less common, our method achieved even higher dosage correlations (*>* 0.98) for all parental phasing scenarios.

We show that the attenuation in the efficacy of embryo screening due to posterior uncertainty is expected to be small for typical embryo cohort sizes when parental haplotypes are accurately estimated, with only a *∼*2% reduction in expected gains when both short and long reads are available for parental phasing.

Large reference panels are becoming increasingly prevalent ^34–36^, but panel composition remains uneven across ancestries. Short-read phasing can suffice for embryo polygenic scoring alone when genetic ancestries are well-matched to the reference panel, but when this is not the case, long reads, grandparental data, more embryos, or higher coverage can compensate. However, some uncertainty in embryo PGS values often remains, highlighting the importance of propagating uncertainty into disease predictions (**Methods**).

Long reads allow us to accurately impute rare variants that may not be present in a reference panel. We illustrate the power of this approach by detecting the presence of a rare pathogenic frameshift mutation in the father of one of the real PGT-A cases analyzed and accurately imputing it in the embryo even when no reads covered the site.

The linear scaling of the ImputePGTA algorithm means it remains computationally tractable even when large numbers of embryos are available, which is increasingly common in contemporary IVF, and which is expected to increase even further with the advent of *in vitro* gametogenesis ^8,37–39^. If large numbers of embryos can be produced using *in vitro* gametogenesis, ULP sequencing followed by imputation could provide a more cost effective approach than producing high coverage data from each embryo. Beyond applications in PGT, the trio-based imputation framework underlying ImputePGTA has broad potential applications, including non-invasive prenatal testing from cell-free fetal DNA, family-based imputation, and refinement of haplotype estimates.

Our method bridges the gap between existing, routine laboratory workflows for PGT-A and the comprehensive genome-wide data required for PGT-P and detection of pathogenic rare variants, lowering the technological barrier to adoption of polygenic embryo testing in assisted reproduction.

## Methods

### The ImputePGTA model

ImputePGTA is a Coupled HMM used to impute offspring genotypes and PGSs from phased parental data and ULP sequence (or genotyping array) data on the offspring. For each informative locus (where an informative locus is defined as those where at least one parent is heterozygous and at least one sequencing read or genotype call is available across all embryos) *l* = 0, 1, …, *L*, the hidden state of offspring *j* = 1, …, *N* is the IV **I**_*j*_[*l*] = (**I**_*pj*_[*l*], **I**_*mj*_[*l*]) *∈ {*(0, 0), (0, 1), (1, 0), (1, 1)*}*, where **I**_*pj*_[*l*] (resp. **I**_*mj*_[*l*]) indexes the paternal (resp. maternal) haplotype whose allele is transmitted. At each position *l*, genotype data (either sequence reads or genotype calls) on embryo *j* = 1, …, *N* is emitted: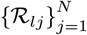. Based on Mendelian laws, the four possible values have equal prior probability, 1*/*4. We make the Markovian assumption that **I**_*j*_[*l* + 1] is conditionally independent of **I**_*j*_ [*l*^*′*^] given **I**_*j*_[*l*] for *l*^*′*^ *< l*.

The true parental haplotypes are denoted by **P** and **M**, and the estimated haplotypes are denoted as 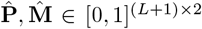. We assume the genotype calls are correct (or that the probabilities are accurate) but that the phasing may be incorrect. To account for switch errors, we introduce a binary master chain **S**[*l*] that encodes the parental switch states at each locus, and which evolves based on parent-specific switch error rates, *λ*_*p*_ and *λ*_*m*_. The offspring IV transitions depend on the master chain, coupling all embryos through shared information about parental switch error locations. This joint modeling allows the algorithm to pool evidence across embryos to more accurately identify and account for switch errors, which is particularly beneficial when parental phasing quality is limited.

We use a Baum–Welch algorithm to obtain maximum likelihood estimates of the paternal and maternal switch-error rates, *λ*_*p*_cM^*−*1^ and *λ*_*m*_cM^*−*1^. The state space scales exponentially in the number of embryos, rendering exact inference via the standard forward-backward algorithm on the full joint model infeasible except for small numbers of embryos. We develop a method that enables us to sample from the joint posterior distribution with computations that scale linearly in the number of embryos, achieving equivalent performance to exact inference.

### Putting *λ* in context

In the literature, switch errors are typically quantified using the switch error *rate* (SER), which is the proportion of pairs of consecutive heterozygotes that are incorrectly phased. Switch errors can be further decomposed into type: a *single switch error* (SSE), in the words of Browning, is a switch error that is not “immediately preceded or followed by another switch error”, whereas a *flip error* (FE) is when two consecutive switch errors occur ^40^. A *phase error* is either a SSE or an FE. For ImputePGTA, it is primarily single switch errors that matter, as these result in long stretches of flipped alleles following a single switch; flip errors manifest merely as a few discordant sites where the flips occurred. Given the sparsity of the embryo data, these double switches are typically undetectable from the embryo data, so *λ* can be interpreted as measuring primarily the number of single switches that occur per cM.

### Expected single switch-error rates in real data

Modern statistical phasing methods applied to the microarray genotypes in the White British cohort in the UK Biobank result in phase error rates equivalent to *λ≈* 0.03 (^40^). However, this represents a best-case scenario, as this estimate derives from common variants on the UK Biobank (UKB) Axiom Array using a reference panel where the ancestry is well-represented. Phasing quality degrades with decreasing minor allele count in the reference panel and for poorly-represented genetic ancestries; as such, statistical phasing performance is highly dependent on the reference panel used ^26,41^. For individuals whose ancestries are not well-represented in readily available reference panels, long read and/or grandparental data can be collected in order to improve phasing ^25^.

### Estimation of polygenic scores

In the context of PGT-P, the statistics of interest for an embryo are their polygenic scores, which are functions of the inheritance vectors (IVs). Under exact inference, the posterior mean PGS can be calculated from the marginal posterior decoding produced by the Forward-Backward algorithm, but this does not give any information on posterior uncertainty in the PGS. However, our inference approach in the joint model enables efficient sampling from the full joint posterior of 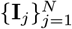 and **S** in *O*(*NL*) time, enabling efficient Monte Carlo estimates of any functional *f* (**I**_*j*_), such as a PGS.

### Prediction of phenotypes and disease risks

Let *Y*_*j*_ be the phenotype of embryo *j* for a quantitative phenotype *Y* . Assuming a simple linear phenotype model based on a PGS, *i.e*., *Y*_*j*_ = *r*PGS_*j*_ + *ϵ*_*j*_, where *ϵ*_*j*_ is an independent error term, the predicted offspring phenotype is a simple linear function of the imputed mean offspring PGS:

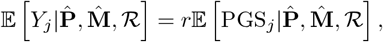

where 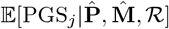 is the posterior mean PGS, which can be computed through posterior decoding under exact inference.

A posterior distribution over the offspring phenotype can be computed by sampling inheritance vectors (above). The target is the conditional density of *Y*_*j*_ given the estimated parental haplotypes and offspring reads:

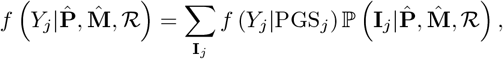

where we have marginalized over the possible offspring inheritance vectors, PGS_*j*_ is a function of 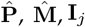 and *f* (*Y*_*j*_|PGS_*j*_) is the density of the phenotype distribution conditional on the PGS, *e.g*., *Y*_*j*_ |PGS_*j*_*∼*𝒩(*r*PGS_*j*_, 1 − *r*^2^). As there are 2^2(*L*+1)^ possible inheritance vectors, it is not practical to evaluate this sum directly. We can instead use samples from the posterior distribution over the inheritance vectors, 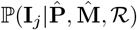, to produce a Monte-Carlo estimate of the posterior predictive distribution. Sample 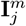 from 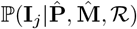 for *m* = 1, …, *M*, then

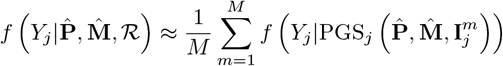

where the PGS for the *m*^th^ sample is computed using the *m*^th^ sampled inheritance vector.

For disease prediction, the probability an offspring develops a disease is usually modelled as a non-linear function of the offspring PGS — for example, using a liability threshold model, potentially including family history and parental PGS — implying that the disease probability is not a linear function of the posterior mean PGS. Let *D*_*j*_ be the event that embryo *j* develops the disease. We assume that 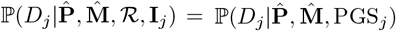, *i.e*., the disease probability model only depends on the inheritance vector and reads through the PGS. Then the posterior probability of disease is

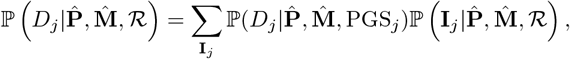

which can be approximated using samples from the posterior distribution of the inheritance vector:

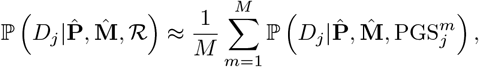

where 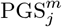 is the PGS computed from the *m*^th^ inheritance vector sampled from the posterior.

### Expected attenuation of selection efficacy when using imputed PGS

The expected gain from embryo selection when the goal is to maximize a quantitative phenotype is the expected difference between the maximum PGS among a number of embryos compared to the average when selecting randomly, scaled by the correlation between PGS and phenotype ^42,43^. When the goal is minimization of disease risk, the expected gain is more complicated to calculate because disease risk is a non-linear function of PGS and can include complex modeling of family history ^43^. However, the expected reduction in PGS value when selecting the embryo with the minimum polygenic risk compared to the average provides a tractable metric that can be interpreted as the reduction in disease liability (as in the standard liability threshold model) when multiplied by the square root of the liability-scale *R*^2^.

The more uncertainty there is in the posterior inheritance vectors, the closer the posterior probability is to 0.5, the unconditional transmission probability based on Mendelian Laws. Thus, posterior uncertainty results in shrinkage of the posterior mean towards the expectation given the parents, which does not vary between embryos and is thus non-informative for embryo selection. We therefore sought to characterize the impact of posterior uncertainty in embryo PGS values on the expected change in true PGS when selecting the embryo with the minimum posterior mean PGS compared to selecting a random embryo.

Here we derive an expression for the expected attenuation of selection efficacy as measured by the relative expected gain when selecting the lowest-risk embryo using posterior mean PGSs compared to when the true value of the PGS is known. We show that the attenuation in the expected gain for selection on a given trait using imputed PGSs is equal to the within-family correlation between the imputed and true PGS, which can be estimated from the posterior and within-family PGS variances.

Assuming normality, Karavani et al. ^43^ show the expected gain from selecting the embryo with the max PGS value, max_*j*_ PGS_*j*_ for *j* = 1, …, *N*, is proportional to the within-family PGS standard deviation (although they do not make this explicit):

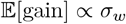

where 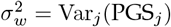 is the within-family variance of the PGS. This is because the expected max of an IID sample from a Normal scales in proportion to the SD.

In the context of ImputePGTA, the PGS of each embryo *j* = 1, …, *N* is estimated by the posterior mean: 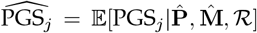, where 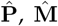 are the estimated parental haplotypes, and *ℛ* represents embryo reads. When using 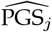 to select an embryo, the gain is equal to the expected true PGS value of the embryo with the maximum imputed PGS value. Since the posterior mean PGS gives the expected value of true PGS for that embryo, the expected gain from selecting the embryo with the maximum imputed PGS value is simply 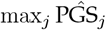. Thus the expected gain is the expected max of the imputed PGS values, which, assuming normality, scales in proportion to the within-family SD of imputed PGS values: 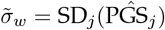.

From the law of total variance, we have that

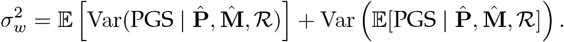

We can write the first term on the RHS as 𝔼[*ν*], which denotes the average variance of the PGS posterior distribution for the embryos, which in practice we can obtain through sampling as described above, and the second term on the RHS is the within-family variance of 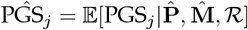. Then we can write 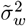 as:

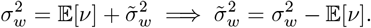

Thus, when selecting on 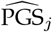, the expected gain is

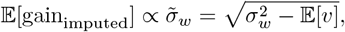

The relative gain that is obtained by selecting on the posterior mean PGS compared to selecting on the true PGS, *i.e*. the *attenuation factor*, can then be written as the ratio of the gains and plugging in the results from above:

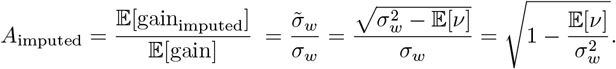

This ratio simplifies to 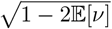 for 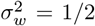, the expected value of within-family PGS variance in a random-mating population with total PGS variance 1.

We now relate this to the within-family correlation between true PGS and imputed PGS:

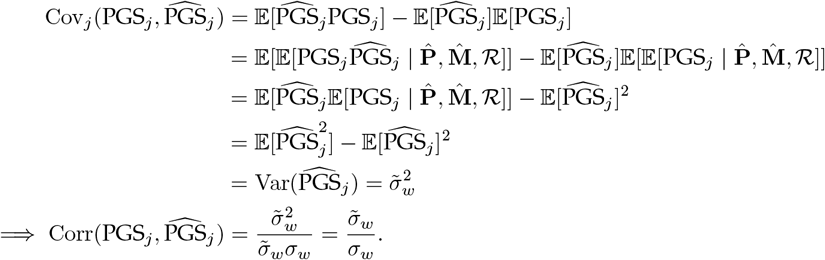

Thus, the attenuation is equal to the within-family correlation between the true and estimated PGS. It is worth noting that while the average within-family PGS variance, 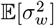, is half the PGS variance in the population under random mating ^44^, the within-family variance for a *particular* family can significantly deviate from this, and is affected by non-random mating.

The within-family variance for a particular family can be estimated by simulation of meiosis. For example, for the Platinum Pedigree, within-family variance calculated from 500 simulated offspring ranged from *∼*80% of the reference population variance for Alzheimer’s disease to *∼*15% for Vitiligo (**Supplementary Table 10**). Small rank-order differences across traits in the empirical gains are within Monte Carlo error (e.g., Breast Cancer 0.923*±*0.040 vs. Alzheimer’s disease 0.900*±*0.038) and reflect sampling noise and departures from Normality, while gains still scale with within-family SD overall.

The PGS for Alzheimer’s disease has an outsized contribution from the APOE locus, implying that there can be substantially more within-family variation than the average for families with one or more parents that are heterozygous for a APOE risk allele (as is the case for the father here). The PGS for vitiligo, in contrast, shows a substantial correlation between parents ^7^ (0.085, S.E. 0.008) — indicative of assortative mating and/or population structure — which is expected to reduce within-family variance as a fraction of total variance. These results show that while theoretical results such as those in Karavani et al. ^43^ give useful estimates of the expected utility of embryo selection at the population level, they rely on assumptions such as random-mating and normality of PGSs that often do not hold in practice, with actual gains for specific diseases in individual families often substantially more or less than expected based on such results.

### *In silico* experiments

A description of the Platinum Pedigree data used for the simulations and in-silico downsampled offspring data is available in **Supplementary Note**. Briefly, we used a family comprising parents (NA12878 and NA12877, both of European ancestry) for which a highly validated phased truth-set comprising SNPs and small indels is publicly available.

#### Simulating offspring and sequencing reads

Simulated offspring genotypes were generated from the phased VCFs for the parents, NA12878 and NA12877, using a genetic recombination model. For each offspring, crossover events were simulated following a Poisson process with rates determined by sex-specific genetic maps ^45^, and offspring genotypes were determined given the transmission of haplotypes implied by the crossover events after first selecting an initial haplotype to transmit. This process was repeated to simulate 20 independent offspring.

Sequencing reads were then simulated from the offspring genotypes using a Poisson sampling process. For each variant site in each simulated offspring, the number of reads was drawn from a Poisson distribution with a rate equal to the target coverage level (ranging from 0.002x to 1x). When reads were present at a site, each read randomly sampled one of the two parental haplotypes with equal probability. Read errors were introduced by randomly changing the true base to one of the other three nucleotides with probability 0.001 (corresponding to Q30 base quality). All reads were assigned fixed Phred-scaled base quality scores of 30, representing high-quality reads.

#### Simulating switch errors in parental haplotypes

Phasing switch errors were introduced into the parental haplotypes using a Poisson process model. For each parent, haplotype data was processed chromosome-wise using sex-specific genetic map positions from the literature ^45^. Switch error events were sampled at each variant position by drawing from a Poisson distribution with a rate equal to the genetic distance (in cM) between consecutive variants multiplied by *λ*, the specified error rate parameter. Here, *λ* can be interpreted as the expected number of switch errors per cM. Switch events occurred when the Poisson draw was odd. Once a switch error was introduced at a given position, all subsequent variants on that chromosome had their haplotype phases inverted until the next switch event occurred, thus modeling the propagation of phasing errors that persist along chromosomal segments. We introduced switch errors at the following rates (*λ*): 0, 0.05, 0.1, 0.2, 0.5, 1.0.

#### Imputation and evaluation of simulated reads and downsampled offspring data

For both the simulated and downsampled experiments, we ran ImputePGTA with either simulated or real reads as the data input for the offspring. As previously described, first, parent-specific switch error rates (*λ*_*p*_, *λ*_*m*_) were estimated from the parental haplotypes, the offspring reads, and a genetic map from the literature ^45^ using a Baum-Welch algorithm; these estimated values were then used in the inference step, where inheritance vectors were estimated and offspring genotypes are imputed.

Imputation accuracy was evaluated using the dosage correlation coefficient as well as genotype concordance. The dosage correlation coefficient is computed as the correlation between the dosage in the imputed VCF and the numeric genotype (*i.e*., 0, 1, 2) in the truth VCF. We also evaluated accuracy at “trio-segregating” (TS), defined as sites where at least one parent is heterozygous, the only sites that vary between siblings other than by *de novo* mutation.

#### Selection efficacy experiment

To characterize the attenuation of expected predicted gains when selecting on the basis of imputed PGS values versus selecting based on true PGSs, we simulated 500 offspring from the Platinum Pedigree parents used in the experiments described above and calculated PGSs for them. We simulated reads at a coverage of 0.004x for each offspring. We then grouped these simulated offspring into 100 batches of 5 embryos each. For each batch, we then simulated estimated parental haplotypes by inducing switch errors at *λ* = 0.05, 0.125, and 1.

The results of this procedure for each value of *λ* represents 100 realizations of embryo batches and estimated parental haplotypes under parameter combinations reflecting the real PGTA cases with ULP sequence data. We used ImputePGTA to jointly impute the offspring genotypes within each family from the simulated read data, and calculated the posterior mean PGS for the 17 traits for each offspring directly from the dosages resulting from posterior decoding. We also drew 500 posterior samples for each imputed offspring and calculated the PGS posterior variance.

We then simulated the process of embryo selection using two strategies: (a) selecting based on the lowest true PGS, and (b) selecting based on the lowest imputed PGS. For each batch of five embryos, we calculated the predicted gain as the difference between the true PGS of the selected embryo and the mean of the true PGSs within the five embryos in the batch. We also calculated the mean posterior variance per trait across all 500 embryos for each *λ* value as well as the true within-family PGS variance to obtain the predicted attenuation factor.

To translate these PGSs onto the risk scale, we used the disease prevalences and liability-scale 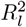 values from Moore et al. 2025^7^ (**Supplementary Tables 11–12**). We used the standard liability threshold model for disease where disease liability *L* is modeled as *L* = *G* + *E* where *G* represents the genetic component distributed as 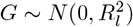 represents the environmental component distributed as 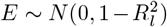, and *G* and *E* are assumed to be uncorrelated. For a binary disease trait with population prevalence *K*, the liability threshold *T* is defined as *T* = Φ^*−*1^(1*− K*) where Φ^*−*1^ is the inverse standard normal cumulative distribution function. The probability of disease (*i.e*., the absolute risk, or AR) for an individual with standardized PGS value *g* is then given by

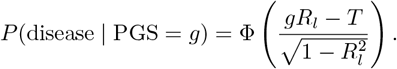

The absolute risk reduction for a given embryo in a batch is then the difference between its absolute risk AR and the mean absolute risk across embryos within the batch 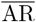. The relative risk reduction (RRR) for a given embryo in a batch of embryos is then 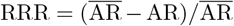. The absolute and relative risk reductions corresponding to the PGS-scale expected gains in **Figure 3** are shown in **Supplementary Figures S3– S4**.

To validate our theoretical result, we simulated 100 batches of 5 offspring genomes from the Platinum Pedigree parents, each of which represents a possible realization of an IVF cycle (**Methods**). For each batch, we induced switch errors in the parents at a rate of *λ* = 0.05cM^*−*1^, 0.125cM^*−*1^, or 1cM^*−*1^, simulated reads from the five offspring at 0.004x coverage, imputed them using ImputePGTA, and drew 100 posterior samples for each of the 17 disease PGSs. These parameters were chosen as they are representative of the empirical results from the real PGT-A cases with ULP data later described. The lower value of *λ* corresponds to results when both long and short reads are used for parental phasing, the second to when only short reads are used, and the third to the highest value of *λ* used in our *in silico* experiments.

For each trait, we then computed the average gain in true PGS across 100 batches when selecting based on the true PGS and on the imputed mean PGS. We also computed the theoretical expected gain as calculated from the posterior variances (**Figure 3**). The translation of these PGSs to absolute and relative risk reductions under a standard liability threshold model are shown in **Supplementary Figures S3 – S4** (see **Methods, Supplementary Tables 11–12**). We found strong agreement between the empirical gain and theoretical gain when selecting on imputed PGS values, indicating that the theoretical result is valid and our posterior variances are well-calibrated. Overall, the attenuation is negligible at *λ* values of 0.05cM^*−*1^and 0.125cM^*−*1^, with *A*_imputed_ = 0.98 and 0.97 respectively, while at *λ* = 1cM^*−*1^, *A*_imputed_ decreased to 0.85, highlighting the importance of accurate parental phasing. The empirical gain when selecting on the true PGS ranged from 0.46 for vitiligo to 0.92 SDs for Breast Cancer, reflecting differences in within-family variance for these PGSs (**Supplementary Table 10**).

### Parental phasing in real data

Statistical phasing of variant calls is generally quite accurate; however, only variants that are present in a reference panel can be phased. An orthogonal approach is read-backed phasing, which uses direct read evidence (such as from long reads) to establish phase, allowing phase estimation at all heterozygotes with sufficient spanning reads ^28^. However, even using modern long read technologies, this results in only multiple disjoint *phase sets* (*i.e*., blocks of phased heterozygotes) that do not span full chromosomes; for example, for one of the real-world cases (FAM1), long read prephasing using high-coverage ONT long reads with whatshap ^28^ on chromosome 1 for the father resulted in phase resolution at >99.9% of all heterozygotes, but contained within 417 disjoint phase sets, meaning that while phase was locally resolvable for these variants, there was insufficient information to link these individual phased blocks together to achieve chromosome-level haplotypes.

Thus, in order to generate chromosome-level haplotypes for the parents that span both statistically phased common variants and long-read-phased rare variants, we used a combination of methods (**Figure 1C**). At a high level, the process is as follows:

1. Mendelian scaffolding (*i.e*., inferring phase using Mendelian laws) using grandparental data (when available) at heterozygotes resolvable by Mendelian inheritance laws.
2. Statistical phasing using `SHAPEIT4`^46^ with the scaffold (when grandparental data was available) option and the phase set option to phase common variants.
3. Read-backed phasing using ONT long reads with `whatshap` to obtain independent phase sets containing both common and rare variants with locally resolved phase.
4. A novel HMM-inspired dynamic programming algorithm we call phaseGrafter to combine the results from (2) and (3) into single unified haplotype estimates spanning both common and rare variants by “grafting” phase sets generated using long reads atop a scaffold of common variants.

For all parents, we used long read data for phasing purposes only, and relied on the short read variant calls for the genotypes. For each sample, we pre-phased and generated phase sets by using Oxford Nanopore long reads `whatshap` v2.6^28^ to phase the short read VCFs, resulting in locally resolved phase sets of heterozygotes.

#### Statistical phasing

We used `SHAPEIT4` v4.2.2 to statistically phase the common variants present in each parent, incorporating Mendelian scaffold and long read phase sets as priors when available.

We used a reference panel of synthetic haplotypes generated using `RESHAPE` ^47^ that is based on the phased reference panel previously generated ^26^ for the UK Biobank 200k WGS release. We subset to variants with MAF *≥*0.1% prior to creating the synthetic haplotypes in order to control computational cost. The result of this is a *common variant scaffold* containing chromosome-level estimated haplotypes, but limited only to common variants overlapping the reference panel used.

#### Rare variant phasing and phaseGrafter

The output of `SHAPEIT4` is a set of statistically phased chromosome level haplotypes for each parent. However, statistical phasing can only resolve variants which are also present in the reference panel used — across the parents of the four families, *∼*9% of heterozygotes remain unresolved in the parents after statistical phasing due to their absence in the reference panel. On the other hand, prephasing has no such limitation, and heterozygotes can be phased where reads span at least two heterozygous variants. The limitation here is instead the disjoint nature of the phase sets, where two independently resolved sets of heterozygotes have internally resolved phase, but the phase relation between them is unclear.

We integrate these two sources of phase information using a novel HMM-inspired dynamic programming approach called phaseGrafter which “stitches” phase sets generated using `whatshap` onto the phased common variant scaffold output from statistical phasing (e.g. `SHAPEIT4`), resulting in unified haplotypes that retain the chromosome-scale consistency of the statistical scaffold at common variants while extending the phasing to rare heterozygotes carried by the individual.

The basic idea behind phaseGrafter is that most variants phased in one dataset will also be phased in the other dataset. For each local haplotype in the read-derived phase sets, one can then compare the relative orientation of the phase estimates in the overlap and deduce the most consistent internal orientation of the heterozygotes within the phase set with respect to the scaffold, after which each heterozygote in the phase set which is *not* in the intersection can be grafted onto the scaffold accordingly.

Using this method, we are able to resolve the phase at *>* 99.9% of heterozygous sites in the parents in the base case (phasing using short and long read data) after applying `phaseGrafter`, thus enabling whole-genome reconstruction of both common and rare inherited variation within offspring. **Supplementary Table 6** contains the variant counts in the parents in addition to the number and percentage that were phased in each scenario.

### Polygenic scores

We calculated PGSs for 17 disease traits described in a recent manuscript from Herasight ^7^: Alzheimer’s disease, atopic dermatitis, basal cell carcinoma, breast cancer, glaucoma, gout, hypertension, inflammatory bowel disease, melanoma, multiple sclerosis, osteoporosis, prostate cancer, psoriasis, testicular cancer, Type 2 diabetes, venous thromboembolism, and vitiligo. These PGS have nonzero weights at *∼*7.3M variants across the autosomes.

To calculate ancestry-adjusted PGS for a given individual, we first computed the PGS for each sample in the HGDP + 1KG dataset ^48^ using the `plink2 --score` function ^49^.

Using these individuals embedded in a common genetic PC space, we then fit per-trait linear models for the PGS conditional mean and residual variance on the first four PCs, then mapped each posterior sample from the target individual into this space and *z*-standardized the raw scores using the predicted mean and variance. The results of this procedure are ancestry-adjusted standardized PGS for each posterior sample.

### Real PGT-A Cases

#### Recruitment of participants and sample collection

We recruited seven families to validate ImputePGTA. The parent participants recruited were informed about the study through their IVF clinic and enrolled under two approved IRB protocols (Solutions IRB protocol ID 0448 and Pearl IRB protocol ID 2025-0250). Informed consent in each case was obtained from both participating parents, and parental consent was obtained on behalf of the born child where applicable. For families where grandparental data was obtained (FAM1 and FAM7), consent was also obtained from the relevant grandparents. **Supplementary Table** 1 shows the details of the families.

For six families (FAM1–FAM6), we obtained PGT-A data from embryos and high-coverage whole-genome sequence data from the corresponding born child(ren) to serve as ground truth. FAM3 and FAM4 each had two born children; the remaining families had one born child each, for a total of eight born children across six families. For the seventh family (FAM7), we obtained high-coverage WGS data from 25 embryos but no data from a born child; instead, we downsampled the WGS data to 0.004x coverage to mimic PGT-A coverages, and used the high-coverage WGS data as truth data for validation.

The parents in all seven families underwent genomic sequencing using both Oxford Nanopore Technologies (ONT) long-read sequencing and Illumina short-read sequencing. The grandparents (if applicable) as well as the born children were sequenced using Illumina short-read sequencing.

To generate long-read data for the parents, whole blood was self-collected at home using the Tasso+ device with an EDTA microtainer tube. Genomic DNA from blood was extracted with the Qiagen Puregene Blood Kit following the manufacturer’s instructions. Libraries were prepared with the Oxford Nanopore Technologies (ONT) Ligation Sequencing Kit and run on a PromethION P2.

To generate short-read data for the born children, grandparents, and parents, self-collection was performed to obtain either buccal epithelial cells, using the iSWAB-DNA-250 kit, or saliva, collected with GeneFiX or Oragene DX kits. For short-read sequencing, libraries were prepared with Illumina DNA Prep and sequenced on an Illumina NovaSeq X+ at Eurofins Clinical Enterprise (Louisville, KY).

The realized short read WGS coverages for aligned and deduplicated reads for the father, mother, and grandparents if applicable are summarized in **Supplementary Table 7**; the average coverage for short reads achieved for the parents and grandparents was 38.9x and the average coverage of the born children/WGS embryo data was 48.3x. For the parental long-read sequencing, the average coverage was 31.4x for the long reads with an average N50 of *∼*31kb.

With assistance from the parents, we obtained the original PGT-A data used for testing during the IVF from the original data providers as described below.

#### Data methods for WGS samples

##### Alignment and variant calling

For the short reads, we aligned them to `GRCh38` (obtained from ftp://ftp.ncbi.nlm.nih.gov/genomes/all/GCA/000/001/405/GCA_000001405.15_GRCh38/seqs_for_alignment_pipelines.ucsc_ids/GCA_000001405.15_GRCh38_no_alt_analysis_set.fna.gz) using `bwa-mem2` v2.3. Long-read sequencing data for the parents were aligned to `GRCh38` using the `epi2me wf_human_variation` workflow v2.6.

The short read alignments were processed using a multi-step variant calling pipeline. For FAM1, FAM2, FAM5, and FAM6 (each with a single born child), we processed the mother, father, and born child using `DeepTrio` v1.9.0 for pedigree-informed variant calling. Joint genotyping of the trio gVCFs was then performed using GLnexus to produce a consolidated trio VCF. All variants were normalized using `bcftools norm` to split multi-allelic sites, filtered to retain only autosomal sites with non-missing genotype calls across all trio members. A further filtering step was done to retain only sites where both parents had confident genotype calls (GQ *≥*30). Individual sample VCFs were extracted from the joint-called trio VCF. For the grandparents in FAM1, we generated single-sample VCFs using `DeepVariant` v1.9.0.

For FAM3 and FAM4 (each with two born children), we generated single-sample gVCFs for the parents and the born children using `DeepVariant` v1.9.0, followed by joint genotyping with GLnexus. The same filters as for the other families were applied.

For FAM7, we jointly called the parents, all four grandparents, and all 25 embryos using `DeepVariant` v1.9.0 followed by joint genotyping with GLnexus. For FAM7, since there was no born child and the sequencing coverage across the pedigree was variable (the mother had *∼*24x coverage), we filtered only on parental genotype quality (GQ *≥*30) without requiring non-missingness across all embryos.

The number of called heterozygous variants passing filters for each parent are tabulated in **Supplementary Table 6**.

##### Embryo PGT-A data

PGT-A data was obtained from the original data provider with assistance from the parents for the embryos corresponding with the born child(ren), as well as any other data from other, unimplanted embryos from the same parents. The data processing steps for embryo ULP sequence and array data are described below.

##### Embryo ULP sequence data

For FAM1, FAM2, and FAM5, the embryo data obtained was ULP sequence data in the form of a BAM file containing single-end reads aligned to `hg19`. For these samples, the PGT-A assay was performed using the Ion ReproSeq PGS Kit (Next Generation Sequencing) for 24 chromosome aneuploidy testing (Thermo Fisher Scientific, USA). The kit/assay was performed on the Ion Chef and Ion S5 System instruments (Thermo Fisher Scientific, Inc, MA, USA). Data analysis was performed with Ion Reporter software (IRv5.16) (Thermo Fisher Scientific, USA).

We reverted the BAMs to FASTQs using samtools and re-mapped the reads using `bwa` v0.7.18 before feeding the reads into ImputePGTA.

After alignment, we mark duplicates using `samtools markdup` and calculate the pileup using `samtools mpileup`, restricted to the set of sites at which at least one parent was variant and reads where the mapping quality (MQ) is *≥*10. We only take into account single bases and thus only SNV variant sites are modeled as having observed data (reads at indels in the parents are ignored). For multi-allelic sites in the parents, we model these as additional states in the HMM separated by a nonzero but negligible genetic distance (in principle, multiallelic variants should be explicitly modeled, but we find that this heuristic works well in practice). These reads after quality score recalibration (described below) are used as the input data for the embryos in ImputePGTA as described in the main text.

#### Embryo genotyping array data processing

For FAM3’s children, the assay used for PGT-A was the Axiom UK Biobank Array ^50^ with *∼*800k SNPs. The data was obtained from the original provider, Genomic Prediction, in the form of VCFs with variant calls on hg19. We lifted these over to GRCh38 using Picard ^51^ and these VCFs were used for input to ImputePGTA. The VCFs obtained from Genomic Prediction did not contain variant-level quality scores.

For FAM4 and FAM6, the assay used for PGT-A was the Illumina HumanCytoSNP-12 BeadChip ^52^ with *∼*300k SNPs. The data was obtained from the original provider, Natera, in the form of IDAT files. We first converted these to GTC format using the Illumina Array Analysis Platform Genotyping Command Line Interface ^53^ followed by conversion to VCF on `GRCh38` using the `gtc2vcf` plugin for `bcftools`^54^.

For each family, the genotype calls in the embryos at variants overlapping with the parental genotype calls are then used as the input to ImputePGTA after quality score recalibration as described below is applied.

We also imputed these arrays to the UKB reference panel used for parental phasing. Briefly, we pre-phased the original VCFs using `SHAPEIT5`^26^, followed by imputation using `IMPUTE5`^55^. We filtered to sites called in the truth VCFs and calculated PGSs using the approach previously described.

##### Base/genotype quality score recalibration

We recalibrated call quality using an embryo-stratified, quantile-based empirical approach. For the sequencing data, Phred-scaled base qualities *Q* were converted to error probabilities as *p* = 10^*−Q/*10^. For array data, we used the genotype qualities (the `FORMAT/IGC` field for IDAT VCF outputs); scores were treated as accuracies and converted as *p* = 1*−* qual. For FAM3’s embryo array data, the VCFs we received did not contain genotype qualities; as such, we assumed a conservative qual = 0.95 for all sites.

Within each embryo, sites were partitioned into five quantiles of the raw quality score; a single stratum was used if the score had zero variance (*e.g*., if quality scores are not available, one could choose to assume a fixed quality score at all sites). At Mendelian-informative sites where both parents were homozygous for the alternate allele, we defined an error as follows. For sequencing, an error was defined as a read base that was not equal to the alt allele at that site. For arrays, an error was defined as a genotype call on the embryo that was not homozygous for the alternate allele. For each embryo-quantile stratum we estimated the observed error rate using Laplace smoothing and compared it with the mean predicted error (based on the uncalibrated scores) in the same stratum.

We adjusted each call’s error probability on the logit scale by adding a shrinkage-weighted offset equal to the difference between observed and predicted logit error rates:

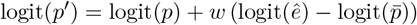

where *w* = *n/*(*n* + *τ*). Here, *p* is the call’s predicted error probability, 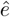 is the stratum’s observed error rate, 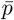 is the stratum’s mean predicted error, *τ* = 30 is a hyperparameter that controls shrinkage toward no adjustment. and *n* is the number of variants in the stratum. When per-embryo strata were sparse or missing, we used quantile-level estimates aggregated across embryos in the same family. Adjusted probabilities were transformed back to the original scales: *Q*^*′*^ = *−*10 *∗*log(*p*^*′*^) for Phred base qualities and qual^*′*^ = 1 *−p*^*′*^ for genotype quality.

##### Evaluation of genotype imputation accuracy

There are regions of the genome known to be technically difficult due to segmental duplications and low-mappability regions, high/low GC regions, tandem repeats, and difficult XY regions. All comparisons were therefore evaluated after applying a custom mask on the genome. Namely, we first mask technically difficult regions of the genome ^56^, defined in the BED file found he re: https://ftp-trace.ncbi.nlm.nih.gov/ReferenceSamples/giab/release/genome-stratifications/v3.5/GRCh38@all/Union/GRCh38_notinalldifficultregions.bed.gz, before adding back any variants in the masked regions that were statistically imputed. In sum, this equates to masking out any rare variants within technically difficult regions. For the scenarios in which we only used short reads for phasing, no sites were masked (*i.e*., all sites that could be statistically phased were used). Across the base case phasing results, an average of 3.3% informative sites across the four families were masked as a result of this procedure.

Genotype imputation accuracy was characterized by (1) the dosage correlation between the imputed dosage at each site and the numeric genotypes in the born child VCFs and (2) genotype concordance, the proportion of imputed hard genotype calls concordant with the genotypes in the born child VCFs. Results are presented primarily for sites that are segregating within the trio (*i.e*., sites where at least one parent is heterozygous), as these are the sites where the inherited genotype is not known *a priori* based on Mendelian inheritance laws.

##### Pathogenic variant in FAM1

A pathogenic variant was detected in the father and child of FAM1. The rare pathogenic frameshift variant in VPS13C (NG_027782.1:g.145304dup, rs1315150327), resulting from a single thymine insertion at chr15:61920162 (`GRCh38`) was detected and called with high confidence in the trio. The child was called heterozygous with a sequencing depth of 51x at the variant and a GQ of 54; the father was called heterozygous with a depth of 47x and a GQ of 44; the mother was called homozygous reference (no mutation) with a depth of 51x and a GQ of 50. In the long read data generated for the father and mother, this site was covered at depths of 38x and 32x respectively. In the PGT-A data for this embryo, this site received no sequencing reads.

## Data Availability

All data from human subjects in this study are not available due to consenting.

## Acknowledgements

We thank Shai Carmi, Joseph Pickrell, Lex Flagel, Robert Maier, Jonathan Anomaly, and Santiago Munne for feedback on an earlier version of the manuscript.

Figure 1 was created using BioRender. This research has been conducted using the UK Biobank Resource under Application Number 103244.

## Author contributions

J.H.L. performed the computational experiments and analyses. A.S.Y. and J.H.L. developed the ImputePGTA model and inference approach; J.H.L. developed and implemented the sampling algorithm; M.C. assisted in the original conceptualization of the method. T.W. implemented the original version of ImputePGTA with assistance from A.S.Y.. A.S.Y. and J.H.L developed extensions to the original mathematical treatments. J.H.L. developed and implemented the phase unification method (phaseGrafter). Sample processing, sequencing, and primary and secondary analyses were validated and performed by J.S. and J.S.. J.H.L., T.W., A.S.Y. designed the study and wrote the manuscript. All authors reviewed the final manuscript. I.D. and J.S. were responsible for setting up the IRB study and provided operational support.

## Supplementary Note: Platinum Pedigree and 1000 Genomes Data Description

### Platinum Pedigree dataset

The Platinum Pedigree resource is a publicly available gold standard pedigree dataset comprising whole genome sequence data from five technologies across a four-generation pedigree ^27^. As of the time of writing this comprises the most fully-validated publicly available dataset for variant calls across multiple generations. We used data from generation 2, which comprises NA12877 and NA12878, the parents, as the source of parental haplotypes for the simulation analyses.

The specific callset we used in this study was the “Pedigree consistent merged small variant calls (truthset)” VCF which comprise a unified callset with pedigree-consistent phased SNPs and small indels for NA12878 and NA12877.

The VCF truthset was obtained from the following s3 path: s3://platinum-pedigree-data/variants/small_variant_truthset/GRCh38/CEPH1463.GRCh38.family-truthset.ov.vcf.gz

We preprocessed the VCF truthset by splitting multiallelics using `bcftools norm -m -any` and filtering to autosomes, resulting in 5949154 variants after deduplication.

### HGDP + 1KG Reference Panel

We used the HGDP + 1KG dataset ^48^, a cosmopolitan dataset representing a diverse set of human genomes, as a reference panel for PGS ancestry adjustment. Briefly, we obtained the raw genotype calls on GRCh38 from the gnomAD website ^31^, annotated them using dbSNP build 157, and filtered them to the variants in our PGS. Ancestry adjustment was then performed as described in the **Methods**.

## Supplementary Figures

**Figure S1.**
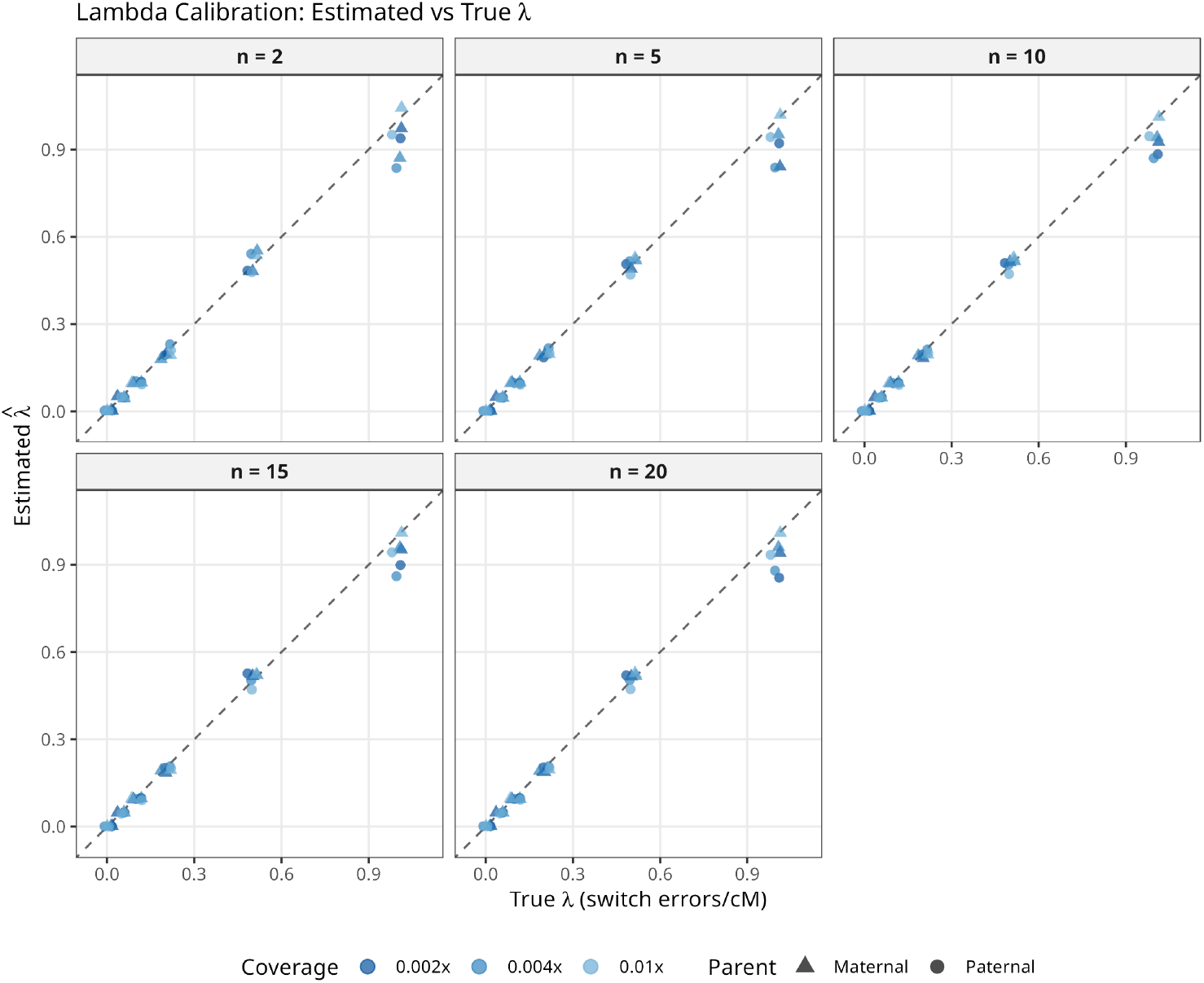
Calibration of lambda estimates. Estimated vs true switch error rates as described by *λ* (switch errors/cM) for the simulated offspring, faceted by the cohort size. The true *λ* reflects the rate at which we introduced switch errors into the parental haplotypes. Points are jittered for visualization. Estimates for the paternal and maternal switch error rates are shown on the *y*-axis and the true simulated *λ* values are shown on the *x*-axis. The estimates are generally accurate and well-calibrated.

**Figure S2.**
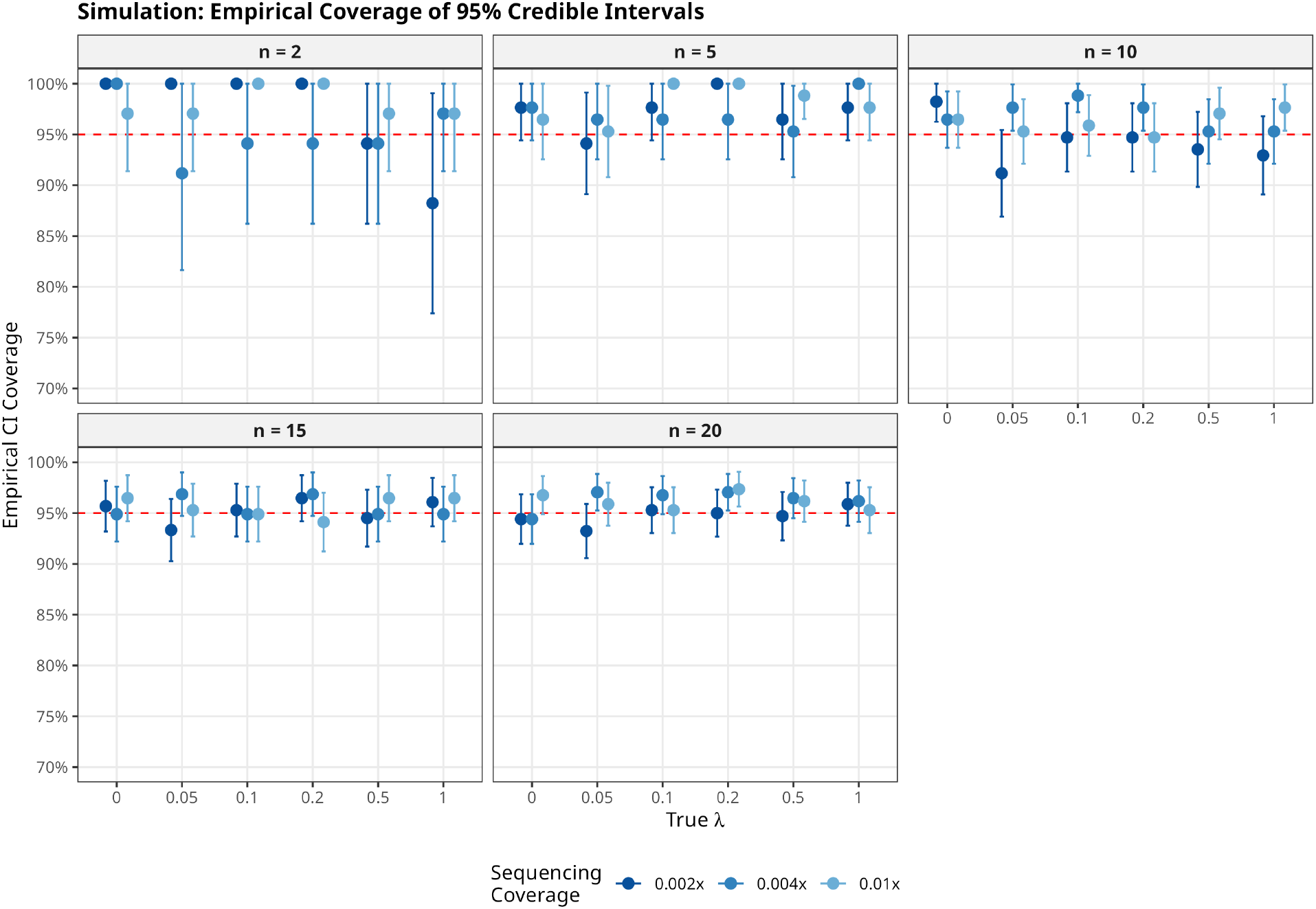
Empirical coverage of PGS posterior 95% credible intervals. The empirical coverage of the 95% equal tailed credible intervals for the in silico experiments. For each coverage x lambda combination, the empirical coverage of the credible intervals across all 5 samples and 17 traits is shown with the standard errors. We observe overall excellent empirical coverage.

**Figure S3.**
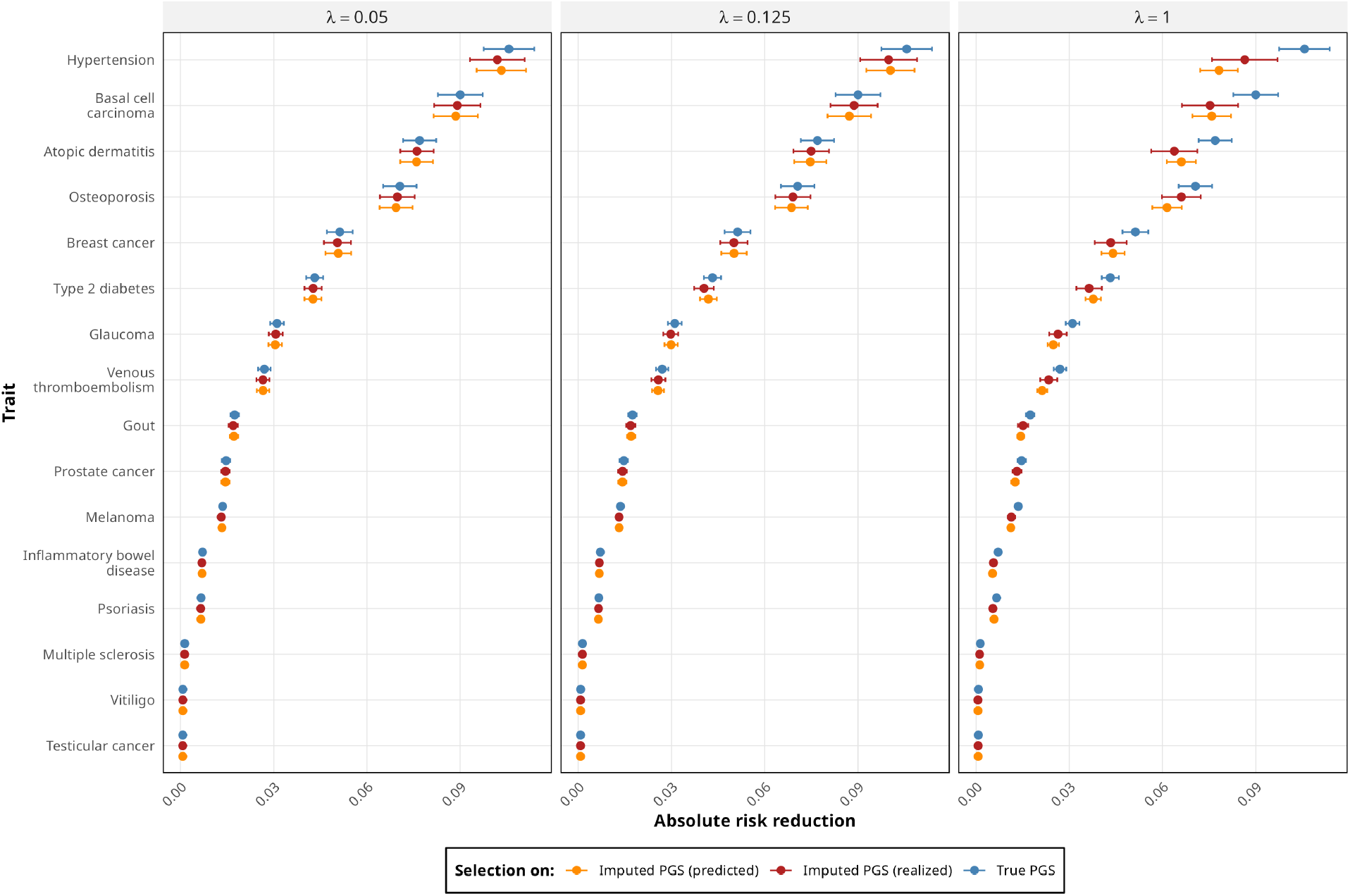
Absolute risk reduction due to selection on imputed PGS. The *x*-axis shows the mean absolute risk reduction (95% CIs) from 100 instances of 5-embryo families with the same parents assuming a liability threshold disease model, where each family’s 5 embryos are jointly imputed. The panels show results for three values of *λ*, the switch-error rate in parental haplotypes: the value based on empirical results for parents phased with short reads and long reads, (*λ* = 0.05), short reads only (*λ* = 0.125), and the highest value analyzed in the *in silico* experiments (*λ* = 1). For each panel, three results are shown: the first two are the absolute risk reductions from two selection strategies: (a) selection based on true PGS values (blue), and (b) selection based on the imputed PGS values (red). The third is the theoretical absolute risk reduction predicted by the posterior variances (orange). The difference between the red and blue data points reflects the actual decrease in absolute risk reduction (relative to true PGS values) resulting from imputation uncertainty.

**Figure S4.**
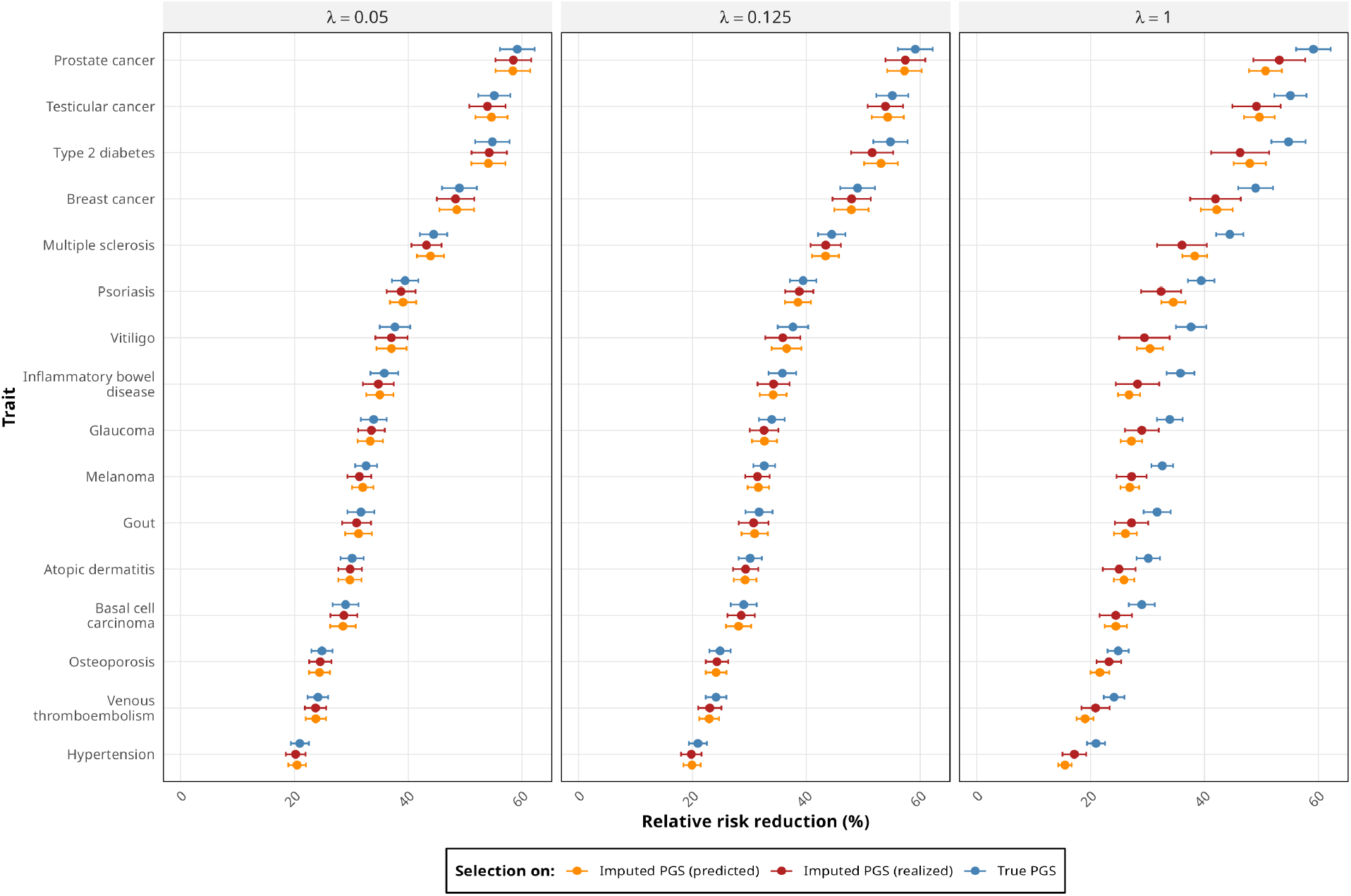
Relative risk reduction due to selection on imputed PGS. The *x*-axis shows the mean relative risk reduction (95% CIs) with respect to the risk corresponding to the within-family embryo mean PGS. These are calculated from 100 instances of 5-embryo families with the same parents assuming a liability threshold disease model, where each family’s 5 embryos are jointly imputed. The panels show results for three values of *λ*, the switch-error rate in parental haplotypes: the value based on empirical results for parents phased with short reads and long reads, (*λ* = 0.05), short reads only (*λ* = 0.125), and the highest value analyzed in the *in silico* experiments (*λ* = 1). For each panel, three results are shown: the first two are the relative risk reductions from two selection strategies: (a) selection based on true PGS values (blue), and (b) selection based on the imputed PGS values (red). The third is the theoretical relative risk reduction predicted by the posterior variances (orange). The difference between the red and blue data points reflects the actual decrease in relative risk reduction (relative to true PGS values) resulting from imputation uncertainty.

**Figure S5.**
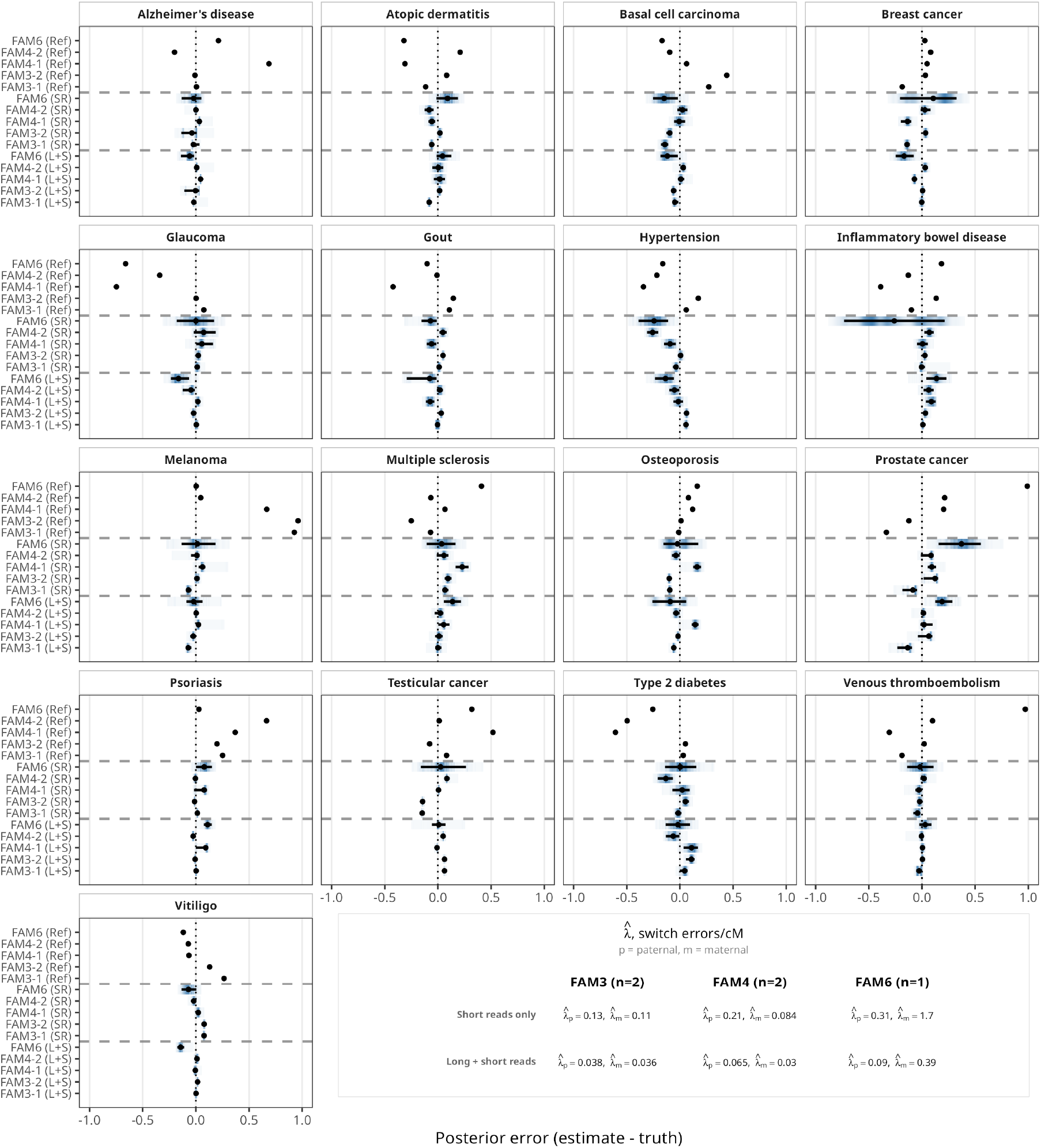
Polygenic score estimates for embryos with genotyping array data. Across 17 disease PGSs, we give posterior PGS estimates in units of standard deviations from the PGS calculated from high-coverage data on the born child. The black points denote the posterior mean PGS, the whiskers denote the 95% equal-tailed credible interval, and the posterior densities are given by the intensity of shading. (SR only) indicates parental haplotypes were produced from short-reads only using statistical phasing. The last four rows show the error of PGS estimates from the results of reference-based imputation. Generally, the CIs are less well calibrated than the ULP cases, but the MAE is substantially lower.

**Figure S6.**
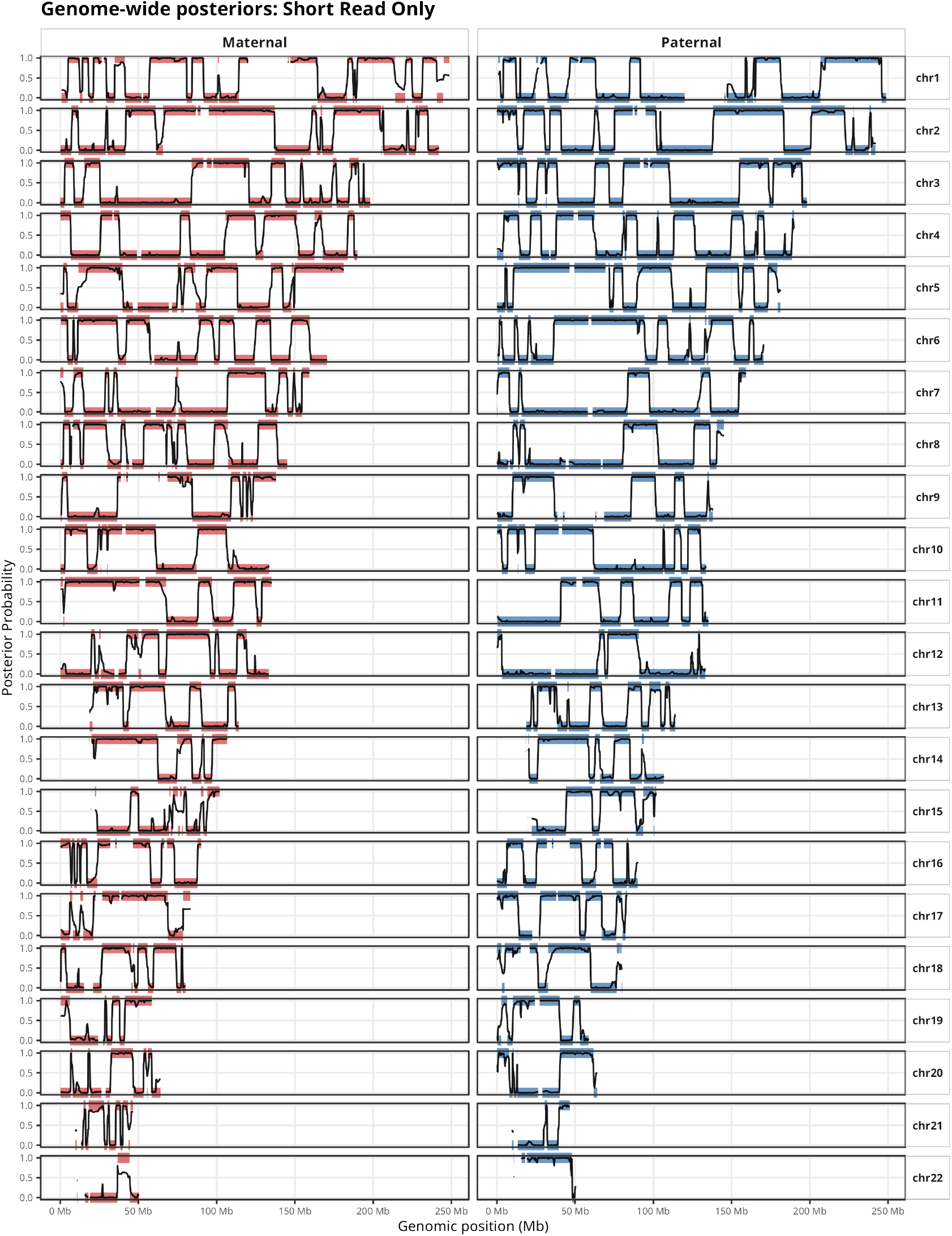
Genome-wide posterior plots for FAM1 parents using short reads only. Each pane depicts in thick colored lines the smoothed posterior probabilities obtained from running ImputePGTA on the born child’s genotypes obtained from high-coverage WGS. Probabilities at 0 and 1 indicate inheritance of one or the other of the parental haplotypes. In black are shown the raw posterior probabilities resulting from running ImputePGTA on the ULP PGT-A data. Gaps on the *x*-axis reflect regions of at least 1Mb where no variants were genotyped in the parents.

**Figure S7.**
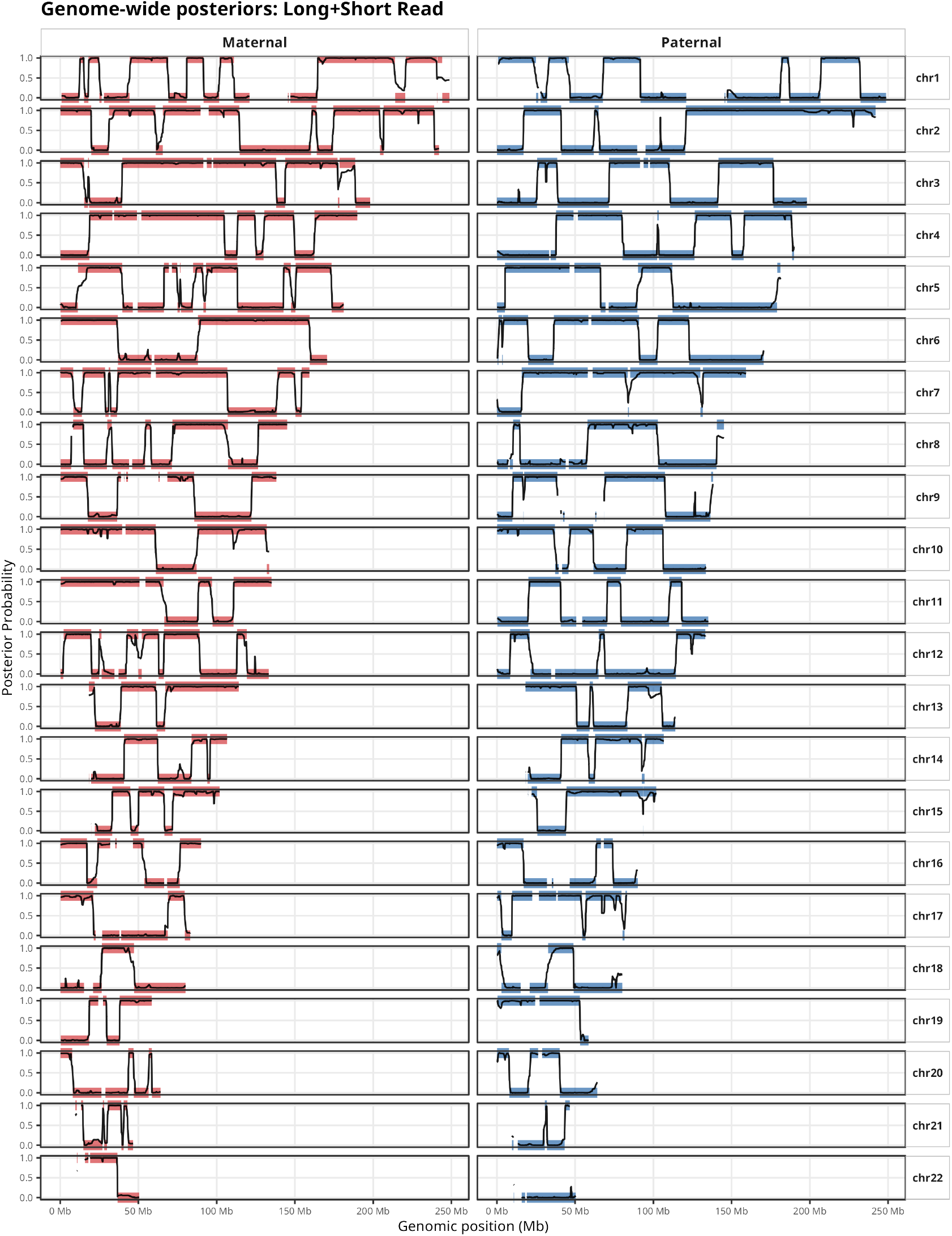
Genome-wide posterior plots for FAM1 parents using short and long reads. Each pane depicts in thick colored lines the smoothed posterior probabilities obtained from running ImputePGTA on the born child’s genotypes obtained from high-coverage WGS. Probabilities at 0 and 1 indicate inheritance of one or the other of the parental haplotypes. In black are shown the raw posterior probabilities resulting from running ImputePGTA on the ULP PGT-A data. Gaps on the *x*-axis reflect regions of at least 1Mb where no variants were genotyped in the parents.

**Figure S8.**
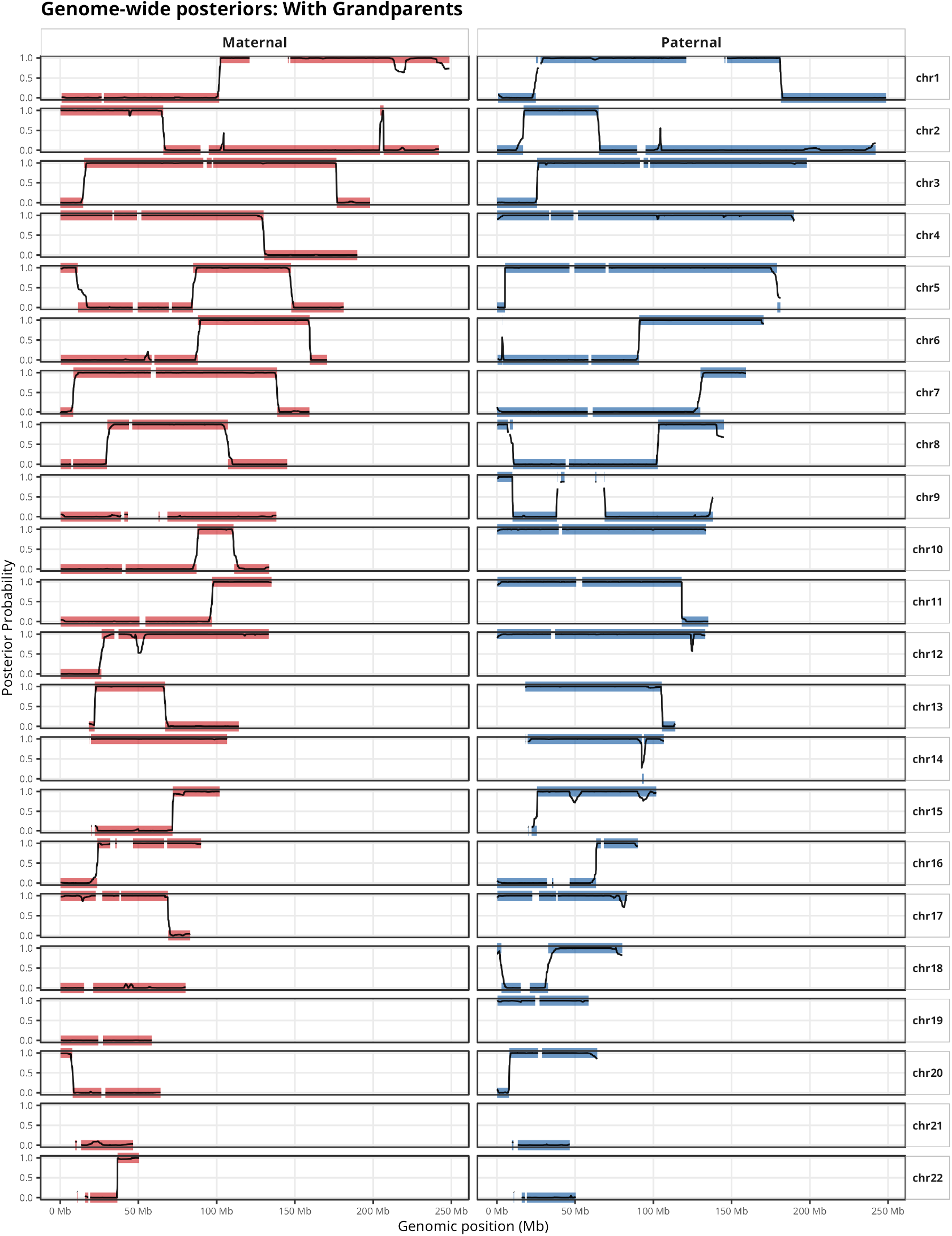
Genome-wide posterior plots for FAM1 parents using short and long reads and grandparental data. Each pane depicts in thick colored lines the smoothed posterior probabilities obtained from running ImputePGTA on the born child’s genotypes obtained from high-coverage WGS. Probabilities at 0 and 1 indicate inheritance of one or the other of the parental haplotypes. In black are shown the raw posterior probabilities resulting from running ImputePGTA on the ULP PGT-A data. Gaps on the *x*-axis reflect regions of at least 1Mb where no variants were genotyped in the parents.

